# No Evidence of Genotype‒Treatment Interactions between Endocrine Therapies and Adverse Drug Effects in Women with Breast Cancer: Findings from the UK Biobank

**DOI:** 10.1101/2025.10.23.25338679

**Authors:** Kinan Mokbel, Michael N. Weedon, Victoria Moye, Katherine S. Ruth, Leigh Jackson

## Abstract

Breast cancer is the most commonly diagnosed cancer worldwide. Earlier studies have demonstrated that breast cancer patients with particular genomic variants are more susceptible to adverse drug effects (ADEs) when they are receiving endocrine therapy. However, to establish a robust body of evidence with regard to the potential utility and predictive value of these variants, findings from these reports require replication. This study aimed to validate previously reported associations between genomic variants and medically important adverse drug effects (MIADEs) using UK Biobank (UKBB). In 2,729 female participants who had received endocrine therapy in the UKBB, this study found no statistically significant interactions between genomic variants and endocrine therapy regarding continuous or binary outcomes. Thus, findings from the UKBB dataset do not support previously documented pharmacogenomic associations of MIADEs in endocrine therapy. In light of current evidence, pharmacogenomic testing within this context should not be considered for individualised endocrine treatment recommendations in clinical practice.

## Introduction

Breast cancer is the most common malignancy worldwide, with approximately 2.3 million new cases diagnosed each year (1). Seventy to eighty per cent of breast cancer cases are hormone receptor-positive (HR+) cases, for which endocrine therapy, including tamoxifen and aromatase inhibitors (AIs), plays a pivotal role in preventing recurrence and improving survival. For HR+ early breast cancer, AIs have shown greater efficacy than Tamoxifen as adjuvant therapy in postmenopausal women, whereas tamoxifen with or without ovarian function suppression is still the appropriate endocrine therapy in premenopausal women (2). Nevertheless, both tamoxifen and AIs significantly reduce relapse rates, increase survival rates and reduce breast cancer mortality when they are administered for a 5–10-year period (3). Breast cancer remains, however, the leading cause of cancer-related death among women, primarily due to metastasis and recurrence (4). Although endocrine therapy has been proven effective for many years, not all women experience its benefits because of a lack of adherence. Endocrine therapy-related adverse drug effects (ADEs), which impact 30–70% of patients, are the main predictors of poor adherence and persistence (5). These symptoms mainly include musculoskeletal, vasomotor, metabolic, vascular, vulvovaginal and endometrial symptoms (6,7). Hence, interventions to improve breast cancer prognosis should encompass measures to prevent the ADEs associated with endocrine therapy.

Emerging evidence suggests that patients with certain genomic variants may be susceptible to clinical toxicity outcomes when undergoing endocrine therapy (8,9), but prior studies have produced conflicting results (10,11). These findings have been marred by substantial heterogeneity and further limited by suboptimal methodological rigour and small sample sizes. Furthermore, the majority of these studies were mainly focused on cohorts of breast cancer patients with specific cancer stages or comorbidities. Thus, it is crucial to establish the reliability of these findings by replicating them successfully in independent and well-designed large cohorts. This study aimed to replicate the previously reported associations between genomic variants and medically important adverse drug effects (MIADEs) associated with endocrine therapy in female participants in the UK Biobank (UKBB) (12). This replication effort is a step forward in determining whether genetic variants need to be considered in clinical practice as a means of preventing MIADEs related to endocrine therapy.

## Materials and Methods

### Description of the Study Population

The UKBB is a cohort of over 500,000 participants from England, Scotland, and Wales, with genomic and health-related data collected from 2006-2010, followed up for up to 14 years, and regularly updated including self-reported data, hospital episode statistics (HES) and linkages to death and cancer registries (13). As the available samples of other ancestries in the UKBB were insufficient in size to draw any reliable conclusions, we limited our analyses to 389,805 unrelated individuals with genetically determined European ancestry using principal component analysis of genomic data, which is distinct from sociopolitical constructs of ethnicity or race. This aligns with the study’s focus on genetic ancestry as a biological construct relevant to pharmacogenomic association analyses. This study was limited to participants with genetically determined European ancestry because of the insufficient sample sizes required for robust statistical analysis among other ancestries represented in the dataset.

We included female participants, defined as individuals who were genetically female and self-reported as female, who self-reported taking endocrine agents at the baseline assessment for the treatment or prevention of recurrence of breast cancer. This comprised endocrine medications including tamoxifen and 3^rd^ generation AIs (letrozole, anastrozole, exemestane). Detailed information on the endocrine agents and other medication codes used in this analysis (Field ID 20003) can be found in Table S1.

### Selection of genomic variants

Owing to the inconsistency and disparate classifications used in the literature to describe and characterise the seriousness of ADEs, we introduced the term MIADEs. With this, we aim to unify the diverse terminology used in the literature to describe the seriousness and severity of ADEs and improve the consistency of reporting, enabling more precise comparisons across different studies. MIADEs include events that are deemed serious or severe by investigators, meet the WHO criteria for seriousness (14,15), are classified as severe as per the Common Terminology Criteria for Adverse Events (CTCAE) (16) or are recognised as designated or important medical events as per the European Medicines Agency (17,18).

To identify genomic variants associated with MIADEs related to endocrine treatment, we curated these variants from our recently published systematic review (9), which involved a comprehensive search across MEDLINE, Embase, Cochrane CENTRAL, Google Scholar and the Pharmacogenomics Knowledge Base (PharmGKB) (19). We applied the specified MIADE criteria and included only variants that showed a statistically significant association with endocrine treatment-related MIADEs.

### Genotyping procedures

The SNP-genotyping array data and imputation used in this study were generated by the UKBB as previously described (12). Stringent quality control procedures were applied by the UKBB team to the dataset (20). We applied additional quality control by including only directly genotyped variants that passed QC or imputed variants with an imputation quality >0.95.

### Ascertainment of biomarkers and phenotypes

For biomarkers and phenotypes indicative of MIADEs, we used data from baseline measurements, linked HES records and follow-up visits. ICD-9/ICD-10 codes and self-reported data were used to create phenotypes. Details on the data field IDs and codes used for ascertainment of baseline measurements and the incident MIADE phenotypes (i.e. after initiation of endocrine therapy or between baseline and follow-up visits) are detailed in Tables S2 & S3. In alignment with some of the initial studies that stratified analyses by menopausal status, we determined menopausal status in our study to ensure replication of their findings. Menopausal status at baseline in the UKBB was determined by self-reported menstrual history: premenopausal women reported regular menses, postmenopausal women had undergone natural or surgical menopause (e.g., hysterectomy or bilateral oophorectomy), as we previously described (21). This yielded three categories: premenopausal, postmenopausal and undefined menopause status. All the data and outcomes are sex specific, as the study population consisted solely of female participants. Thus, sex-based comparisons were not applicable, and outcomes were not assessed for sex-specific differences.

### Data analysis and statistical methods

We used linear regression to examine associations between baseline measurements and genetic variants, and logistic regression for associations between incident MIADE phenotypes and genetic variants. This involved two distinct analyses: one for continuous outcomes using baseline data, and another for binary outcomes using baseline data, follow-up visits, and updated HES.

Regression analyses using main effects and interaction effects models were used. The main effects model was employed to assess the risk of MIADEs by comparing individuals with and without a specific genotype, and multivariate regression was used to control for confounding factors. Additionally, we introduced interaction terms into the models to evaluate whether the treatment modified the relationship between genotype and MIADE risk. Unlike main effects modelling, which assumes linear and additive effects, the inclusion of interaction terms (treatment × genotype) in the regression model allows for the detection of non-additive effects, indicating whether the impact of genotype on MIADE risk is modified by treatment.

Variants were analysed using dominant, recessive, or additive genetic models, as specified in the initial studies. Statistical analyses were adjusted for the assessment centre and the first five genetic principal components to account for population stratification, and potential confounders, as reported in the initial studies. Bonferroni correction was used to account for multiple testing, resulting in a critical *p-value* of 5.15E-4 (0.05 corrected for 97 tests). STATA (version 16.0) was used for data management and statistical analysis, and the analyses were two-tailed.

### Ethical approval

UKBB was approved by the UKBB Research Ethics Committee as well as the Northwest Multicentre Research Ethics Committee (MREC). All participants provided informed consent at the time of enrolment. Our access to UKBB data was under application numbers (49847 and 9072).

## Results

### Twenty-four studies reported associations between variants and MIADEs

We identified 41 genomic variants significantly associated with MIADEs related to endocrine treatment across 24 studies (22,23,32–41,24,42–45,25–31) (Table 1). These studies were categorised by system organ class, treatment modality and number of variants examined (Table 2). Musculoskeletal and reproductive MIADEs were the most studied outcomes, examined in 42% and 21% of the studies, respectively (Figure S1). The 41 variants were identified in 19 genes, with *CYP19A1*, *ESR1* and *CYP2D6* being the most frequently examined, accounting for 27%, 15% and 12% of the total number of analysed variants, respectively (Figure S2). Notably, only 12.5% of the studies appropriately incorporated statistical interactions, effect modifications or interaction effects models in their analyses (Table S4).

**Table 1.**
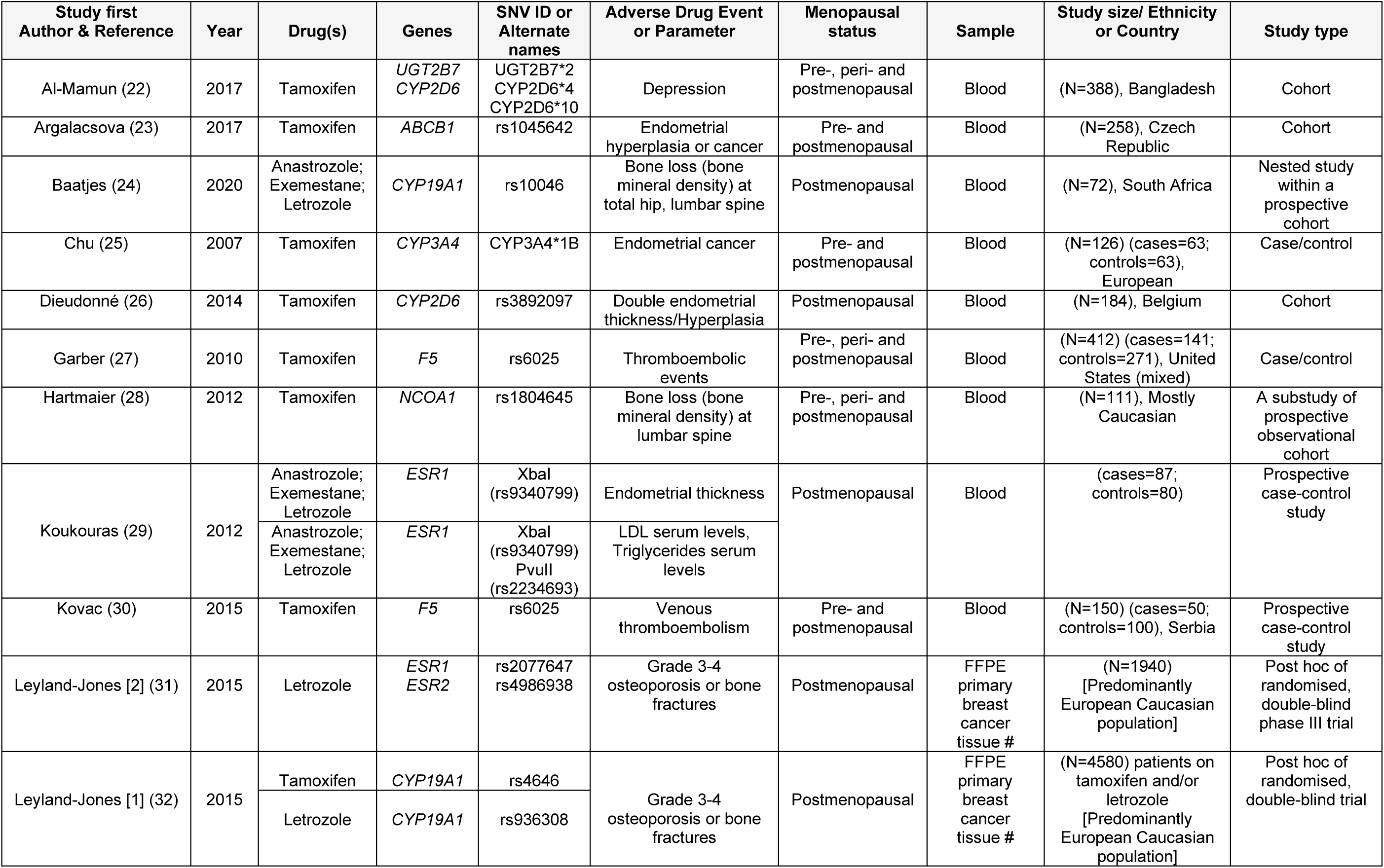
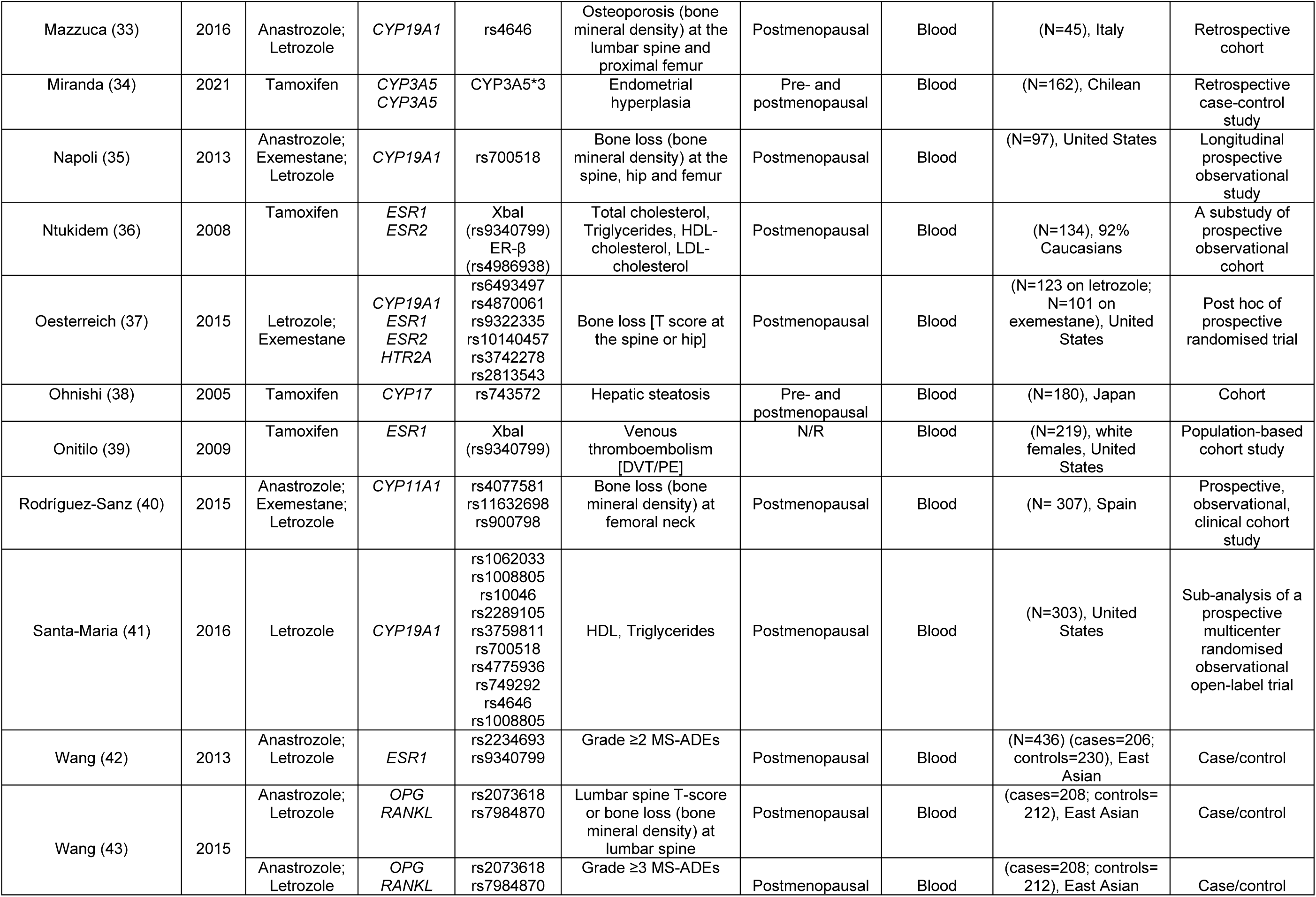
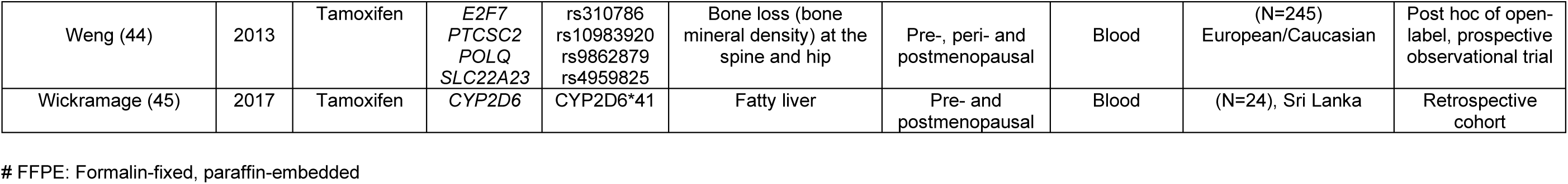
Main characteristics of the studies included in this analysis.

**Table 2.** Studies included in this analysis grouped by system organ class, treatment modality, number of SNVs and related MIADEs.

| System Organ Class | Endocrine agent & variants (n) | Adverse Effects | Studies (n) | Reference. |
| --- | --- | --- | --- | --- |
| Musculoskeletal Disorders | Tamoxifen (n=7),<br>Aromatase Inhibitors (n=18) | BMD*, T-score*, Bone fractures, Osteoporosis*, MS-ADEs | (n=10) | (24,28,32,33,35,37,40,42–44) |
| Metabolism Disorders | Tamoxifen (n=2),<br>Aromatase Inhibitors (n=11) | Hypercholesterolaemia*, Hypertriglyceridaemia* | (n=3) | (29,36,41) |
| Hepatobiliary Disorders | Tamoxifen (n=4) | Hepatosteatosi | (n=2) | (38,45) |
| Vascular Disorders | Tamoxifen (n=3) | Thromboembolic events (incl. DVT, PE) | (n=3) | (27,30,39) |
| Reproductive System Disorders | Tamoxifen (n=4),<br>Aromatase Inhibitors (n=1) | Endometrial cancer,<br>Endometrial Hyperplasia | (n=5) | (23,25,26,29,34) |
| Psychiatric Disorders | Tamoxifen (n=3) | Depression | (n=1) | (22) |

\* **Continuous measurements** or **binary outcomes** derived from baseline measurements.

### 2,729 women in the UK Biobank were receiving endocrine therapy

In the UKBB, we identified 2,729 female participants who received endocrine therapy (mean age=59.2 years). Of these, 1,195 were on tamoxifen (271 premenopausal and 825 postmenopausal) and 1,544 were on AIs (59 premenopausal and 1,261 postmenopausal). Among the AI group, 1,016 were on Anastrozole, 312 were on letrozole, and 218 were on exemestane (Figure 1). The participant characteristics are detailed in Table 3.

**Figure 1.**
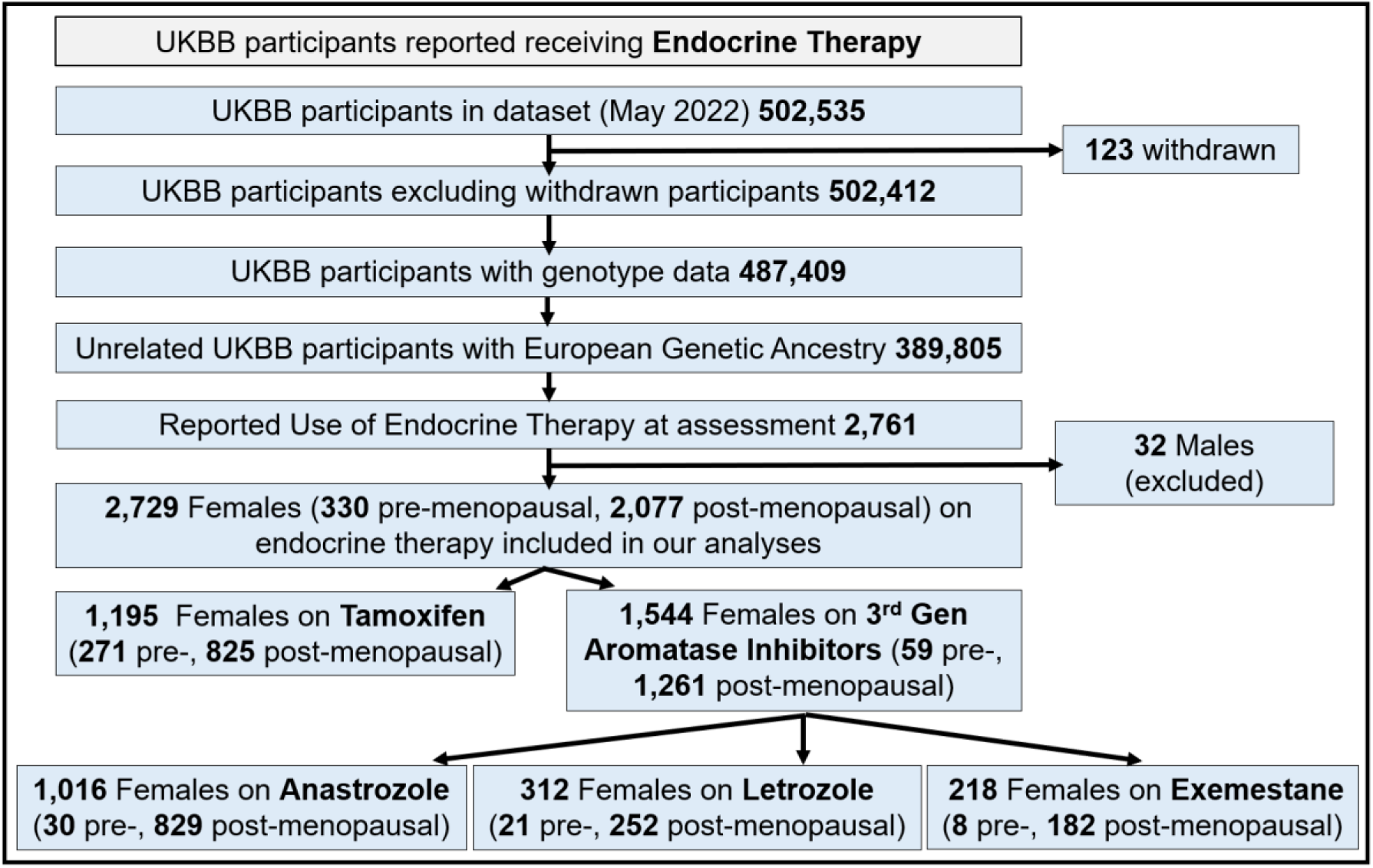
The UK Biobank cohort of patients who reported taking endocrine agents. A flow chart demonstrating the number of UKBB female participants with sufficient genomic and treatment data included in the analyses.

**Table 3.** Characteristics of the UK Biobank female participants taking endocrine agents.

|  | Treatment | Endocrine therapy |  |  |  | The rest of the UKBB female Participants |  |  |  |
| --- | --- | --- | --- | --- | --- | --- | --- | --- | --- |
| Main characteristics | Menopausal Status | Premenopausal (n=330) | Postmenopausal (n=2,077) | Other (n=322) | Total (n=2,729) | Premenopausal (n=55,819) | Postmenopausal (n=132,736) | Other (n=19,164) | Total (n=207,719) |
|  | Age (years) Mean (SD) | 49.6 (5.1) | 60.2 (6.2) | 62.7 (5.1) | 59.2 (7) | 47.4 (4.5) | 60.2 (5.7) | 62.6 (5.2) | 57 (7.9) |
|  | BMI (kg/m<sup>2</sup>) Mean (SD) | 25.7 (4.4) | 27.4 (4.9) | 27.7 (5.1) | 27.2 (4.9) | 26.4 (5.3) | 27.1 (5) | 27.6 (5.2) | 27 (5.1) |
| Continuous outcomes | Bone Mineral Density (g/cm<sup>2</sup>) Mean (SD) | 0.51 (0.11) | 0.49 (0.11) | 0.48 (0.1) | 0.49 (0.11) | 0.55 (0.12) | 0.5 (0.12) | 0.5 (0.12) | 0.52 (0.12) |
|  | T-score Mean (SD) | -0.58 (0.97) | -0.83 (1.02) | -0.96 (0.88) | -0.82 (1) | -0.24 (1.04) | -0.68 (1.06) | -0.73 (1.06) | -0.57 (1.07) |
|  | Total cholesterol [mmol/L] Mean (SD) | 5.3(1.03) | 5.81 (1.12) | 5.89 (1.19) | 5.76 (1.13) | 5.45 (0.97) | 6.07 (1.13) | 6 (1.17) | 5.9 (1.12) |
|  | Triglycerides [mmol/L] Mean (SD) | 1.53 (0.92) | 1.77 (0.97) | 1.84 (1.02) | 1.75 (0.98) | 1.3 (0.74) | 1.64 (0.87) | 1.7 (0.88) | 1.55 (0.85) |
|  | LDL [mmol/L] Mean (SD) | 3.13 (0.79) | 3.57 (0.87) | 3.61 (0.92) | 3.52 (0.88) | 3.33 (0.76) | 3.76 (0.87) | 3.71 (0.91) | 3.64 (0.87) |
|  | HDL [mmol/L] Mean (SD) | 1.59 (0.39) | 1.57 (0.37) | 1.55 (0.39) | 1.57 (0.38) | 1.56 (0.36) | 1.62 (0.38) | 1.59 (0.39) | 1.6 (0.38) |
| Binary outcomes | Osteoporosis [n (%)] | 4 (1.31) | 56 (3.03) | 2 (0.7) | 62 (2.54) | 177 (0.36) | 2,452 (2.06) | 442 (2.58) | 3,071 (1.65) |
|  | Bone fractures [n (%)] | 31 (9.84) | 138 (7.04) | 34 (11.22) | 203 (7.88) | 1,935 (3.59) | 8,727 (6.92) | 1,577 (8.71) | 12,239 (6.17) |
|  | MS-ADEs [n (%)] | 0 (0) | 2 (0.1) | 1 (0.31) | 3 (0.11) | 21 (0.04) | 115 (0.09) | 17 (0.09) | 153 (0.07) |
|  | Hepatosteatosi s [n (%)] | 4 (1.21) | 49 (2.37) | 5 (1.56) | 58 (2.13) | 542 (0.97) | 2,113 (1.6) | 390 (2.04) | 3,045 (1.47) |
|  | Thromboembolic events [n (%)] | 7 (2.16) | 70 (3.54) | 22 (7.33) | 99 (3.8) | 502 (0.91) | 2,871 (2.24) | 528 (2.86) | 3,901 (1.93) |
|  | Venous thromboembolism (DVT/PE) [n (%)] | 6 (1.85) | 48 (2.42) | 12 (3.99) | 66 (2.53) | 311 (0.57) | 1,706 (1.33) | 288 (1.55) | 2,305 (1.14) |
|  | Endometrial cancer [n (%)] | 4 (1.21) | 27 (1.3) | 4 (1.25) | 35 (1.29) | 183 (0.33) | 784 (0.59) | 109 (0.57) | 1,076 (0.52) |
|  | Endometrial hyperplasia [n (%)] | 10 (3.05) | 8 (0.39) | 1 (0.31) | 19 (0.7) | 203 (0.36) | 241 (0.18) | 45 (0.24) | 489 (0.24) |
|  | Depression [n (%)] | 14 (4.62) | 84 (4.41) | 19 (6.83) | 117 (4.7) | 1,934 (3.78) | 4,808 (3.91) | 1,003 (5.77) | 7,745 (4.04) |

\*Values are presented as the mean (SD) [range min-max] or [number of cases (%)]

### No previously reported genotype‒treatment interactions replicated in the UK Biobank

In UKBB, 44% of the variants reported by the investigators of the 24 initial studies were directly genotyped, and 56% were imputed with high confidence (>95%) (Table 4). The initial studies investigating the pharmacogenomics of MIADEs related to endocrine treatment reported 97 statistically significant associations, including 46 associations for continuous outcomes and 51 for binary outcomes (Figure 2).

**Figure 2.**
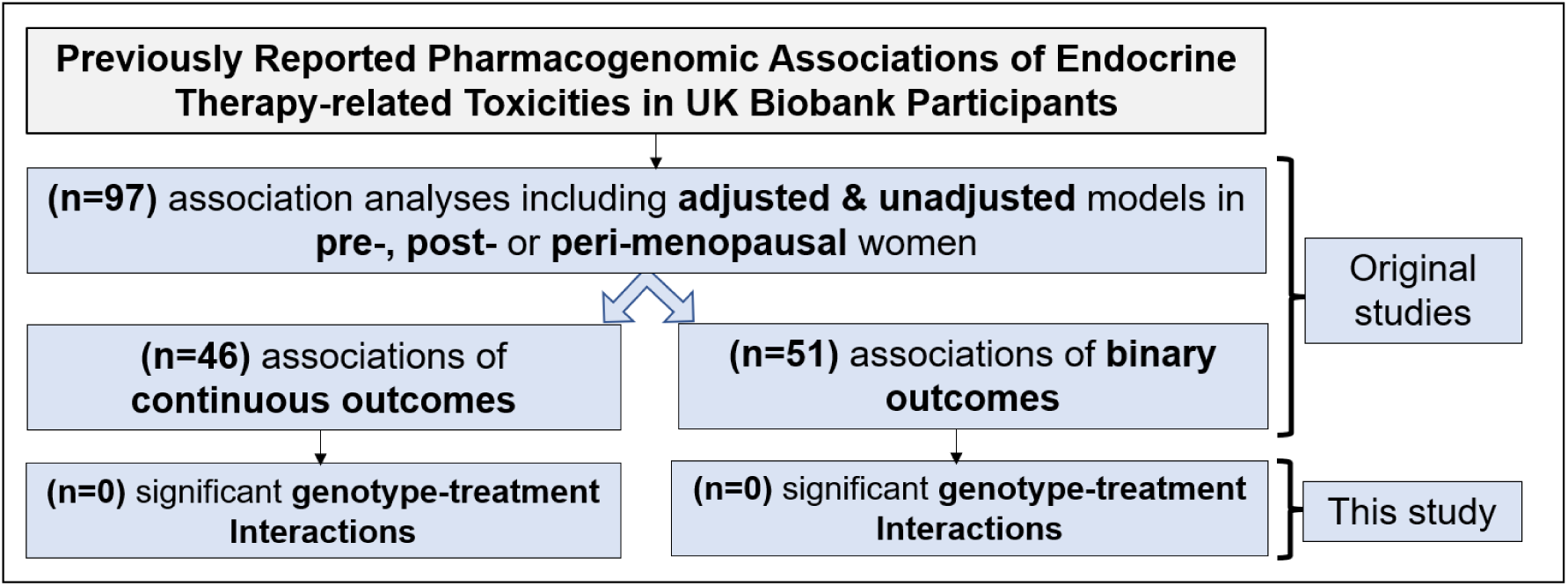
The main results from the UK Biobank analysis of PGx of endocrine therapy-related MIADEs. Associations between previously reported SNVs and MIADEs related to endocrine therapy were tested in UKBB participants. No statistically significant interactions between treatment and mutation status for the risk of MIADEs for any of the variants analysed were observed.

**Table 4.** The genomic variants analysed including frequencies of reference and minor alleles.

| Gene | SNV ID | Variant type/Consequence | Directly genotyped or Imputed | Imputation Score R2* | Chromosome number | Position** | allele 1 | allele 2 | Minor Allele UKBB | MAF UKBB (Unrelated Europeans) |
| --- | --- | --- | --- | --- | --- | --- | --- | --- | --- | --- |
| CYP19A1 | rs10046 | 3 Prime UTR | Genotyped | N/A | 15 | 51502986 | G | A | G | 0.47 |
|  | rs1008805 | Intronic | Imputed | 0.990 | 15 | 51549599 | G | A | G | 0.42 |
|  | rs1062033 | Intronic | Imputed | 0.991 | 15 | 51547938 | C | G | G | 0.46 |
|  | rs3759811 | Intronic | Imputed | 1 | 15 | 51529265 | T | C | T | 0.49 |
|  | rs4646 | 3 Prime UTR | Genotyped | N/A | 15 | 51502844 | A | C | A | 0.26 |
|  | rs4775936 | Intronic | Imputed | 0.996 | 15 | 51536022 | C | T | T | 0.48 |
|  | rs2289105 | Intronic | Imputed | 0.997 | 15 | 51507508 | T | C | T | 0.47 |
|  | rs6493497 | Upstream | Genotyped | N/A | 15 | 51630835 | G | A | A | 0.12 |
|  | rs700518 | Synonymous | Genotyped | N/A | 15 | 51529112 | T | C | T | 0.49 |
|  | rs936308 | Intronic | Imputed | 0.994 | 15 | 51581074 | C | G | G | 0.14 |
|  | rs749292 | Intronic | Imputed | 0.997 | 15 | 51558731 | G | A | A | 0.45 |
| CYP11A1 | rs4077581 | Promoter | Imputed | 1 | 15 | 74665514 | C | T | C | 0.30 |
|  | rs11632698 | Intronic | Imputed | 0.998 | 15 | 74637867 | A | G | A | 0.38 |
|  | rs900798 | 3 Prime UTR | Imputed | 0.995 | 15 | 74629070 | T | G | T | 0.31 |
| CYP2D6 | rs1065852 | Missense | Imputed | 0.994 | 22 | 42526694 | G | A | A | 0.22 |
|  | rs1080985 | Upstream | Imputed | 0.991 | 22 | 42528382 | C | G | C | 0.23 |
|  | rs16947 | Missense | Imputed | 0.997 | 22 | 42523943 | A | G | A | 0.33 |
|  | rs3892097 | Splice Acceptor | Imputed | 0.992 | 22 | 42524947 | C | T | T | 0.21 |
|  | rs28371725 | Intronic | Imputed | 0.989 | 22 | 42523805 | C | T | T | 0.10 |
| ESR1 | rs9322335 | Intronic | Imputed | 0.977 | 6 | 152200129 | T | C | T | 0.26 |
|  | rs9340799<br>(XbaI) | Intronic | Genotyped | N/A | 6 | 152163381 | A | G | G | 0.35 |
|  | rs2077647 | Synonymous | Genotyped | N/A | 6 | 152129077 | T | C | C | 0.48 |
|  | rs2234693<br>(PvuII) | Intronic | Genotyped | N/A | 6 | 152163335 | T | C | C | 0.46 |
|  | rs2813543 | Intronic | Imputed | 0.957 | 6 | 152424478 | A | G | A | 0.23 |
|  | rs4870061 | Intronic | Imputed | 0.997 | 6 | 152237468 | T | C | T | 0.25 |
| ESR2 | rs10140457 | Intronic | Imputed | 0.994 | 14 | 64716693 | A | C | C | 0.02 |
|  | rs4986938 | Non-coding | Genotyped | N/A | 14 | 64699816 | C | T | T | 0.38 |
| F2 | rs1799963 (F2<br>FII G20210A) | 3 Prime UTR | Imputed | 0.950 | 11 | 46761055 | G | A | A | 0.01 |
| F5 | rs6025 (FVL) | Missense | Genotyped | N/A | 1 | 169519049 | T | C | T | 0.02 |
| CYP17A1 | rs743572 | 5 Prime UTR | Genotyped | N/A | 10 | 104597152 | A | G | G | 0.38 |
| CYP3A4 | rs2740574 | Upstream | Genotyped | N/A | 7 | 99382096 | C | T | C | 0.03 |
| CYP3A5 | rs776746 | Splice Acceptor | Genotyped | N/A | 7 | 99270539 | C | T | T | 0.07 |
| TNFRSF11B | rs2073618 | Missense | Genotyped | N/A | 8 | 119964052 | G | C | C | 0.45 |
| TNFSF11 | rs7984870 | Intronic | Imputed | 0.997 | 13 | 43146482 | G | C | C | 0.45 |
| PTCSC2 | rs10983920 | Intronic | Imputed | 0.997 | 9 | 100602613 | C | A | A | 0.12 |
| NCOA1 | rs1804645 | Missense | Genotyped | N/A | 2 | 24974958 | C | T | T | 0.03 |
| E2F7 | rs310786 | Intronic | Genotyped | N/A | 12 | 77436148 | C | T | C | 0.14 |
| *ABCB1* | rs1045642 | Missense | Genotyped | N/A | 7 | 87138645 | A | G | G | 0.46 |
| *SLC22A23* | rs4959825 | Intronic | Imputed | 0.988 | 6 | 3412240 | T | C | T | 0.31 |
| *UGT2B7* | rs7439366 | Missense | Genotyped | N/A | 4 | 69964338 | T | C | C | 0.46 |
| *POLQ* | rs9862879 | Downstream | Genotyped | N/A | 3 | 121149009 | C | T | T | 0.12 |

\*R<sup>2</sup> is the squared correlation between input genotypes and imputed dosages (i.e., true and inferred genotypes)
\*\*Genomic position, build 37 (hg19)

There were a few significant associations in the main effects model. Tamoxifen-treated women carrying the Factor V Leiden (FVL) variant in the *F5* gene had increased odds of venous thromboembolism and thromboembolic events under the unadjusted dominant model [OR (95% CI), *P*]: 1.40 (1.18, 1.66), 9.1E-05 and 1.62 (1.43, 1.83), 5.60E-14, respectively. However, the genotype‒treatment interaction for these events was not statistically significant: [OR (95% CI), *P*]: 3.02 (1.09, 8.33), 0.033 and 1.95 (0.78, 4.88), 0.15, respectively. In the main effects model, women with prothrombotic mutations (FVL or FII G20210A) also showed increased odds of venous thromboembolism (DVT/PE) in the main effects model [OR (95% CI), *P*]: 1.54 (1.35, 1.77), 4.10E-10], but this was not significant in the interaction model [OR (95% CI), *P*]: 2.88 (1.18, 7.03), 0.02. The sensitivity analysis indicated that this effect was driven primarily by FVL, as the interaction between FII G20210A and treatment for DVT/PE was not statistically significant according to either the unadjusted or adjusted models: [OR (95% CI), *P*]: 2.09 (0.46, 9.42), 0.34 and 3.04 (0.65, 14.25), 0.16, respectively.

In the main effects model, postmenopausal women on 3rd Gen AIs carrying the rs700518 *CYP19A1* variant showed a significant association with lower BMD (β, 95% CI, P: -0.003 [-0.005, -0.002], 4.54E-06), but this association was not statistically significant in the interaction model (β, 95% CI, *P*: 0.005 [-0.011, 0.020], 0.55). Similarly, those with the rs10046 *CYP19A1* variant had lower odds of >5% bone loss [OR (95% CI), *P*: 0.95 (0.93, 0.97), 6.80E-05], but this was not statistically significant in the interaction model [OR (95% CI), *P*: 0.97 (0.73, 1.28), 0.82]. For the rs2073618 *TNFRSF11B* variant, postmenopausal women taking anastrozole or letrozole had higher odds of osteopenia [OR (95% CI), *P*: 1.07 (1.04, 1.10), 6.70E-07], but this did not hold in the interaction model [OR (95% CI), *P*: 1.18 (0.86, 1.62), 0.31].

Across all 97 regression analyses, no statistically significant interactions were observed between treatment and mutation status for either continuous or binary outcomes (Figure 2). A statistically significant interaction would imply that the relationship between the independent and dependent variables is modified by a third variable, such as how endocrine treatment modulates the impact of a genomic variant on MIADEs. Although the main associations were replicated, the effect sizes were similar regardless of whether the participants received endocrine treatment. All association analyses are detailed in Tables S5 & S6.

## Discussion

Previous research has shown that genomic variants may affect toxicity outcomes in breast cancer patients receiving endocrine treatment (22,23,32–41,24,42–45,25–31). However, methodological limitations, small sample sizes and conflicting findings underscore the importance of replicating these findings in large, independent cohorts to ensure robustness.

In this extensive investigation of a large population cohort, none of the previously reported associations were replicated for either continuous or binary outcomes. These findings contradict those reported in the initial studies, underscoring the importance of conducting large-scale and independent cohort replication prior to considering pharmacogenomic variants in clinical practice. Our findings are however consistent with PharmGKB’s current low evidence level (Level 3) assigned to these associations, reflecting the lack of consistent replication across studies. Notably, many previous investigations failed to identify significant associations between the variants and endocrine therapy-related ADEs despite the large number of tests performed and in spite of examining multiple variants and several toxicity endpoints (46,47,56–65,48,66–75,49,76,77,50–55), suggesting potential false-positive findings.

Potential implications for practice and research include the importance of exercising caution when interpreting findings from pharmacogenomic studies and the necessity for adherence to rigorous methodological practices. In contrast to this study, most initial studies failed to consider genotype‒treatment interactions in their analyses, which were incorporated into our regression models to minimise bias. Only 12.5% of the initial studies appropriately incorporated statistical interactions in their analyses. In contrast to this study, which carefully adjusted for covariates and applied Bonferroni correction to manage multiplicity, some earlier studies may not have consistently incorporated these adjustments, potentially increasing the risk of false positives and less robust conclusions (78). The differences between our findings and those of previous studies suggest that not accounting for interaction effects and multiplicity may have impacted earlier results.

While the study performed an extensive analysis using a large cohort with longitudinal data that is significantly longer than the initial studies and adhered to rigorous methodological practices, it is important to acknowledge a few limitations. First, the phenotypic data within the UKBB, especially those self-reported by participants at baseline, might vary in reliability and quality, which could pose challenges in accurately identifying individuals with relevant conditions (79). Second, since this analysis included UKBB participants with European ancestry, genetic ancestry findings from this study are not applicable to non-Caucasian populations or broader racial and ethnic groups. Third, participants within the UKBB cohort who carry risk variants might be generally healthier than carriers in the general population, potentially attenuating the pharmacogenomic effects (80). However, our findings are less confounded compared to those from traditional clinical trials since UKBB participants were not informed about their possession of any specific genetic variant. Finally, while controlling for multiple tests is crucial for minimising Type I errors, it also increases the risk of Type II errors (overlooking true positive effects). This issue is further amplified in large datasets such as the UKBB dataset, where background noise can obscure true associations. Thus, smaller and more targeted studies may detect significant results for specific genotype‒drug‒ toxicity interactions that our study did not identify.

In conclusion, this study is the largest to date in its attempt to replicate pharmacogenomic associations between previously identified variants and MIADEs in breast cancer patients undergoing endocrine treatment. None of the previously reported associations were replicated, indicating a lack of evidence for their predictive value and, consequently, their potential use in this context for individualised endocrine therapy recommendations in breast cancer. Thus, alternative interventions may be necessary to manage endocrine therapy-related MIADEs in patients with breast cancer. There is a need for both genomic and non-genomic biomarkers to predict endocrine treatment-related MIADEs. Other approaches, including GWASs using datasets from diverse populations with extended follow-up periods, hold the potential for identifying novel genomic safety biomarkers.

## Acknowledgements

This study received support from the University of Exeter Sanctuary Scholarship. We extend our gratitude to the UK Biobank for granting access to their data for this research. For the purpose of open access, the author has applied a Creative Commons Attribution (CC BY) licence to any Author Accepted Manuscript version arising from this submission.

## Author Contributions

KM, MW, VM and LJ were responsible for designing the research. KM generated the data, performed the analyses, interpreted the results, created the tables/figures, searched the literature and drafted the manuscript. LJ, MW and VM oversaw the study, provided expert interpretation of the data and contributed to the manuscript. KR contributed to data generation and the manuscript. All the authors contributed to the final paper.

## Data availability statement

The genetic and phenotypic data from the UK Biobank can be accessed by applying through their website (https://www.ukbiobank.ac.uk/register-apply). We cannot directly grant access to the specific data fields used in this study. All other data relevant to the study are included in the article or uploaded as supplementary information.

