## Supplemental Files for "No Evidence of Genotype‒Treatment Interactions between Endocrine Therapies and Adverse Drug Effects in Women with Breast Cancer: Findings from the UK Biobank"

### Supplementary Materials

*Table S1 Endocrine agents and other medications codes used in the UK Biobank analysis.*

| Treatment modality | Generic name | Brand names | Codes* |
|---|---|---|---|
| Endocrine Therapy | Tamoxifen | TAMOXIFEN, EMBLON, NOLTAM, NOLVADEX, Soltamox, tamofen | 1140870164, 1140870182, 1140858348, 1140870170, 1140870176, 1141170264 |
| | Letrozole | LETROZOLE, FEMARA | 1141145896, 1141145900 |
| | Anastrozole | ANASTROZOLE, ARIMIDEX | 1140923018, 1140923022 |
| | Exemestane | EXEMESTANE, AROMASIN | 1141171100, 1141171104 |
| Bone Antiresorptive Therapy | Alendronate sodium | Fosamax, Binosto, Fosavance, Bentexo | 1140922174, 1141176570 |
| | Ibandronic acid | Bondronat, Iasibon, Bonviva, Quodixor | 1141180314, 1141190534 |
| | Pamidronate disodium | | 1140868784 |
| | Risedronate sodium | Actonel | 1141175684, 1141175690 |
| | Zoledronic acid | Zometa, Aclasta, Zerlinda | 1141173814 |
| | Teriparatide | Forsteo, Movymia, Terrosa | 1141188794, 1141188798 |
| | Raloxifene | Evista | 1141168574, 1141168578 |
| | Denosumab | Prolia, Xgeva | N/A |
| | Romosozumab | Evenity | N/A |
| Lipid-lowering Therapy | Lipid-lowering drug (generic) | | 1140861922 |
| | Simvastatin | Zocor, zocor heart-pro, INEGY | 1140861958, 1140881748, 1141200040 |
| | Pravastatin | Lipostat | 1140888648, 1140861970 |
| | Rosuvastatin | Crestor | 1141192410, 1141192414 |
| | Fluvastatin | Lescol | 1140888594, 1140864592 |
| | Atorvastatin | Lipitor | 1141146234, 1141146138 |
| | Ezetimibe | Ezetrol | 1141192736, 1141192740 |
| | Acipimox | Olbetam | 1140861892, 1140861894 |
| | Bezafibrate/Bezafibrate product | Bezalip, Fibrazate | 1140861924, 1141157260, 1141201306, 1140861926, 1140861928 |
| | Ciprofibrate | | 1140862026 |
| | Fenofibrate | Supralip, Lipantil | 1140861954, 1141172214, 1141162544 |
| | Gemfibrozil/Gemfibrozil product | Lopid | 1140861856, 1141157262, 1140861858 |
| | Colestipol | Colestid | 1140888590, 1140861848 |
| | Cholestyramine/Cholestyramine product | Questran | 1140909780, 1141180722, 1141180734, 1140861936 |
| | Clofibrate | | 1140861944 |

*Treatments and medication codes (including those used as used as covariates) were obtained from:
https://biobank.ctsu.ox.ac.uk/crystal/coding.cgi?id=4&nl=1
https://biobank.ndph.ox.ac.uk/showcase/field.cgi?id=20003

*Table S2 Ascertainment of biomarkers and other phenotypes at the baseline*

##### A. Cholesterol, LDL, HDL and Triglycerides biomarkers at the baseline

| Description | Data-Field | Items of Data/Participants | Units of measurement | Measurement method |
|---|---|---|---|---|
| Cholesterol | 30690 | 487,377/470,756 | mmol/L | Measured by CHO-POD analysis on a Beckman Coulter AU5800 |
| LDL direct | 30780 | 486,454/469,918 | mmol/L | Measured by enzymatic protective selection analysis on a Beckman Coulter AU5800 |
| HDL cholesterol | 30760 | 429,793/429,793 | mmol/L | Measured by enzyme immunoinhibition analysis on a Beckman Coulter AU5800 |

| | | | | |
|---|---|---|---|---|
| Triglycerides | 30870 | 486,969/470,386 | mmol/L | Measured by GPO-POD analysis on a Beckman Coulter AU5800 |

#### B. Bone Mineral Denisty, T-score, Osteoporosis and Osteopenia Variables at the baseline

##### *1) Bone Mineral Denisty*

Bone Mineral Density (BMD) g/cm2. The Heel bone mineral density (BMD):
https://biobank.ndph.ox.ac.uk/ukb/field.cgi?id=3148
Method: Heel ultrasound method (left). Bone-densitometry of heel:
https://biobank.ndph.ox.ac.uk/ukb/field.cgi?id=4092
This uses BMD and if it is missing, we used Heel bone mineral density (BMD), manual entry:
https://biobank.ndph.ox.ac.uk/ukb/field.cgi?id=3084
And if this was missing, we used Heel bone mineral density (BMD) (left) or Heel bone mineral density (BMD) (right):
https://biobank.ndph.ox.ac.uk/ukb/field.cgi?id=4105
https://biobank.ndph.ox.ac.uk/ukb/field.cgi?id=4124

The following data Field IDs were used for **BMD**:
3148 Heel bone mineral density (BMD)
4105 Heel bone mineral density (BMD) (left)
4124 Heel bone mineral density (BMD) (right)
3084 Heel bone mineral density (BMD), manual entry

##### *2) T-score*

**T-score** (number of SD BMD above or below standard) was computed from **Heel bone mineral density (BMD) T-score, automated** (bone-densitometry of heel). The following data Field IDs were used:
78 Heel bone mineral density (BMD) T-score, automated
4125 Heel bone mineral density (BMD) T-score, automated (right)
4106 Heel bone mineral density (BMD) T-score, automated (left)
4138 Heel bone mineral density (BMD) T-score, manual entry (left)
4143 Heel bone mineral density (BMD) T-score, manual entry (right)

##### *3) Osteoporosis and Osteopenia*

**Osteoporosis and Osteopenia** were calculated based on **Heel bone mineral density (BMD) T-score** as follows:
Osteopenia: T score less than or equal to -1 but greater than -2.5
Osteoporosis: T score less than or equal to -2.5

*Table S3 Ascertainment of incident phenotypic endpoints of adverse drug effects and other variables used in the UK Biobank analysis.*

**A. Incident phenotypic endpoints of adverse drug effects and other phenotypes used as co-variates**

| Phenotype | ICD10 | ICD9 | Self-report code from n_20002_* variable |
|---|---|---|---|
| **Scoliosis (including Kyphoscoliosis)** | M41 M965 Q763 Q675 | 7373 75420 | 1535 |
| **Osteoporosis** | M80 M81 M82 | 7330 | 1309 |
| **Fractures** | M484 M840 M841 M842 M843 M844 S02 S12 S220 S222 S223 S224 S228 S229 S32 S42 S52 S62 S72 S82 S92 T02 T08 T10 T12 T14 M800 M801 M804 M805 M808 M809 | 80 81 82 7331 E8879 | 1626 1627 1628 1629 1630 1631 1632 1633 1634 1635 1636 1637 1638 1639 1640 1644 1645 1646 1647 1648 1649 1650 1651 1652 1653 1654 1655 1656 |
| **Severe musculoskeletal adverse events (e.g., myopathy, myositis)** | G720 G728 G729 M608 M609 M814 | 3599 | 1322 |
| **Thromboembolic events** | I82 I26 I801 I802 I803 I808 I809 I81 I82 I630 I631 I633 I634 I74 K550 I82 I513 I676 I636 I240 | 453 4151 4341 444 59381 3259 4340 4376 4511 4512 4518 4519 4529 59382 | 1094 1093 1088 1068 |
| **Venous thromboembolism (DVT/PE)** | I82 I26 I513 I676 I636 I240 | 453 4151 3259 4340 4376 4511 4512 4518 4519 4529 59382 | 1094 1093 1068 |
| **Hepatosteatosis** | K76 K760 K758 | 5715 5716 5718 5719 | |
| **Gynaecological events [endometrial thickening, uterine fibroids, ovarian cysts, adenomyosis and cervical canal cysts]** | N850 N851 D25 N830 N832 N800 Q516 | 6213 2191 2360 218 6200 6201 6202 | 1351 1349 |
| **Gynaecological events [endometrial hyperplasia or endometrial cancer]** | N850 N851 C541 | 6213 2191 2360 182 | 1040 |
| **Endometrial hyperplasia or Double endometrial thickness** | N850 N851 | 6213 2191 2360 | |
| **Depression** | F33 F32 F412 F341 | 3004 311 | 1286 |
| **Varicose veins** | I83 I86 | 454 456 | 1494 |
| **Endometrial cancer** | C541 | 182 | 1040 |
| **Prosthetic limb by prosthesis** | Z441 Z971 | V437 V520 | |
| **Chemotherapy** | Z082 Z511 Z542 Z926 | V581 V662 V6621 V6622 V6629 V672 | |
| **Radiotherapy** | Z081 Z091 Z510 | V580 V661 V6611 V6612 V6619 V671 | |
| **Family history of ischaemic heart disease** | Z824 | V173 | |
| **Family history of stroke** | Z823 | V171 | |
| **Family history of musculoskeletal diseases or arthritis** | Z826 | V177 V178 | |
| **Family history of malignant neoplasm of genital organs** | Z804 | V164 | |
| **Family history of genital disease** | Z804 Z842 | V187 V164 | |

| | | |
|---|---|---|
| **Family history of psychiatric and other mental and behavioural disorders** | Z818 | V170 |
| **Family history of blood disease** | Z832 | V183 |

#### B. Family history variables of cancer, depression or stroke used as co-variates

Data field IDs and codes for relevant illnesses of father/mother/siblings were used

**Field ID for illnesses of father/mother/siblings:**
20107 Illnesses of father
20110 Illnesses of mother
20111 Illnesses of siblings

**Coding for illnesses of father/mother/siblings:**
2 Stroke
3 Lung cancer
4 Bowel cancer
5 Breast cancer
13 Prostate cancer
12 Severe depression

#### C. Menopausal status

**Menopausal status** was determined at the baseline UK Biobank visit:
**0=premenopausal**
**1=postmenopausal due to natural menopause**
**2=postmenopausal due to surgery i.e., hysterectomy or oophorectomy**
**3=postmenopausal but can't tell what type of menopause or age at menopause as taking HRT over age at menopause**
**4=premenopausal but taking HRT so can't be sure**
9=missing data for one or more variables so can't define this.

*Table S4 Status of including interaction effects in analytical models conducted by authors of pharmacogenetic studies on MIADEs related to endocrine therapy.*

| Study [Author, Year] | The interaction effects model was used appropriately* |
|---|---|
| Al-Mamun 2017 | No |
| Argalacsova 2017 | No |
| Baatjes 2020 | No |
| Chu 2007 | No |
| Dieudonné 2014 | No |
| Garber 2010 | No |
| Hartmaier 2012 | No |
| Koukouras 2012 | No |
| Kovac 2015 | No |

| | |
|---|---|
| Leyland-Jones 2015 (1) | Yes |
| Leyland-Jones 2015 (2) | Yes |
| Mazzuca 2016 | No |
| Miranda 2021 | No |
| Napoli 2013 | No |
| Ntukidem 2008 | No |
| Oesterreich 2015 | No |
| Ohnishi 2005 | No |
| Onitilo 2009 | No |
| Rodríguez-Sanz 2015 | No |
| Santa-Maria 2016 | Yes |
| Wang 2013 | No |
| Wang 2015 | No |
| Weng 2013 | No |
| Wickramage 2017 | No |

**The frequency of the failure to consider potential interaction effects in the analyses was assessed using the following search terms: moderator, heterogeneity of effect, magnifier, qualifier, synergy, buffering effect, statistical interactions, effect modification, effect modifier, multiplicative or synergistic.*

### Supplementary Table S5

| System Organ Class | Regimen or Drug(s) | Gene | SNP rs# ID | Genotype | Statistical genetic model | Toxicity endpoint or Parameter | Menopausal status | Original study | | | Replication attempt in UK Biobank | | | | | |
|---|---|---|---|---|---|---|---|---|---|---|---|---|---|---|---|---|
| | | | | | | | | Original study | Adjustement | Effect size | Adjustment | Statistical model | Effect size | IC95% | UC95% | p_value |
| **Musculoskeletal Disorders** | **Anastrozole; Exemestane; Letrozole** | ***CYP19A1*** | **rs700518** | **A** | **Recessive** | **Bone mineral density** | **Post-menopausal** | **Napoli 2013** | **BMI and baseline BMD** | **Means % Change BMD with 1 y± SD: AA vs. AG+GG [-2.684± 2.34 vs. -0.4± 2.11], (P = 0.03)** | **BMI, assessment centre, first five principal components** | **Main effects model** | **β coefficient=-0.003** | **-0.005** | **-0.002** | **4.54E-06** |
| | | | | | | | | | | | | Interaction model | β coefficient=0.00477239 | -0.010756 | 0.0203008 | 5.47E-01 |
| Musculoskeletal Disorders | Tamoxifen | *NCOA1* | rs1804645 | T | Additive | Bone mineral density | Pre- , Peri- & Post-menopausal | Hartmaier 2012 | N/R (individuals receiving chemotherapy were excluded from analysis) | Mean % Change BMD with 1 yr of tamoxifen therapy in the lumbar (WT:−1.30% ± 0.55, n=107; SNP:−6.43% ± 1.62, n=4; P=0.037) | Assessment centre, first five principal components [those receiving chemotherapy were excluded] | Main effects model | β coefficient=-0.00120996 | -0.0035125 | 0.0010925 | 3.03E-01 |
| | | | | | | | | | | | | Interaction model | β coefficient=0.00848767 | -0.0309076 | 0.047883 | 6.73E-01 |
| Musculoskeletal Disorders | Letrozole; Exemestane | *CYP19A1* | rs6493497 | A | Recessive | T-score | Post-menopausal | Oesterreich 2015 | Age, BMI, prior chemotherapy or tamoxifen | Mean Change T score (SD)= VT/VT −2.90 (0) vs. WT/WT, WT/VT −0.37 (0.44), p=4.6E−8 | Age, BMI, prior chemotherapy, assessment centre, first five principal components | Main effects model | β coefficient=-0.0119326 | -0.063995 | 0.0401298 | 6.50E-01 |
| | | | | | | | | | | | | Interaction model | β coefficient=-0.4482673 | -1.630686 | 0.7341511 | 4.60E-01 |
| Musculoskeletal Disorders | Letrozole; Exemestane | *ESR1* | rs4870061 | T | Recessive | T-score | Post-menopausal | Oesterreich 2015 | Age, BMI, prior chemotherapy or tamoxifen | Mean Change T score (SD)= VT/VT −1.07 (1.01) vs. WT/WT, WT/VT −0.34 (0.40), p=3.2E−7 | Age, BMI, prior chemotherapy, assessment centre, first five principal components | Main effects model | β coefficient=-0.0138345 | -0.0383734 | 0.0107045 | 2.70E-01 |
| | | | | | | | | | | | | Interaction model | β coefficient=0.1409758 | -0.2913455 | 0.5732971 | 5.20E-01 |

| | | | | | | | | | | | | | | | | |
|---|---|---|---|---|---|---|---|---|---|---|---|---|---|---|---|---|
| Musculoskeletal Disorders | Letrozole; Exemestane | *ESR1* | rs4870061 | T | Recessive | T-score | Post-menopausal | Oesterreich 2015 | Age, BMI, prior chemotherapy or tamoxifen | Mean Change T score (SD)=WT/VT −0.39 (0.40) vs. VT/VT 1.07 (1.01) vs. WT/WT −0.31 (0.40), p=5.2E−5 | Age, BMI, prior chemotherapy, assessment centre, first five principal components | Main effects model | β coefficient=-0.0089843 | -0.0187114 | 0.0007429 | 7.00E-02 |
| | | | | | | | | | | | | Interaction model | β coefficient=0.1186838 | -0.0598134 | 0.297181 | 1.90E-01 |
| Musculoskeletal Disorders | Letrozole; Exemestane | *ESR1* | rs9322335 | T | Recessive | T-score | Post-menopausal | Oesterreich 2015 | Age, BMI, prior chemotherapy or tamoxifen | Mean Change T score (SD)=VT/VT 0.44 (0.63) vs. WT/WT, WT/VT −0.29 (0.47), p=3.5E−5 | Age, BMI, prior chemotherapy, assessment centre, first five principal components | Main effects model | β coefficient=-0.0031943 | -0.0271939 | 0.0208054 | 7.90E-01 |
| | | | | | | | | | | | | Interaction model | β coefficient=0.4719414 | 0.0120968 | 0.931786 | 4.40E-02 |
| Musculoskeletal Disorders | Letrozole | *ESR1* | rs4870061 | T | Recessive | T-score | Post-menopausal | Oesterreich 2015 | Age, BMI, prior chemotherapy or tamoxifen | Mean Change T score (SD)=VT/VT −1.25 (0.90) vs. WT/WT, WT/VT −0.36 (0.39), p=3.1E−6 | Age, BMI, prior chemotherapy, assessment centre, first five principal components | Main effects model | β coefficient=-0.0138436 | -0.0383835 | 0.0106962 | 2.70E-01 |
| | | | | | | | | | | | | Interaction model | β coefficient=0.1247933 | -0.4244687 | 0.6740553 | 6.60E-01 |
| Musculoskeletal Disorders | Letrozole | *ESR1* | rs9322335 | T | Recessive | T-score | Post-menopausal | Oesterreich 2015 | Age, BMI, prior chemotherapy or tamoxifen | Mean Change T score (SD)=VT/VT 0.54 (0.63) vs. WT/WT, WT/VT −0.35 (0.40), p=5.2E−6 | Age, BMI, prior chemotherapy, assessment centre, first five principal components | Main effects model | β coefficient=-0.0031724 | -0.0271728 | 0.0208281 | 8.00E-01 |
| | | | | | | | | | | | | Interaction model | β coefficient=0.5900829 | -0.0185096 | 1.198675 | 5.70E-02 |
| Musculoskeletal Disorders | Exemestane | *CYP19A1* | rs6493497 | A | Recessive | T-score | Post-menopausal | Oesterreich 2015 | Age, BMI, prior chemotherapy or tamoxifen | Mean Change T score (SD)=VT/VT −2.90 (0) vs. WT/WT, WT/VT −0.30 (0.38), p=1.6E−9 | Age, BMI, prior chemotherapy, assessment centre, first five principal components | Main effects model | β coefficient=-0.0115203 | -0.0635841 | 0.0405434 | 6.60E-01 |
| | | | | | | | | | | | | Interaction model | β coefficient=-0.4287698 | -1.617906 | 0.7603667 | 4.80E-01 |
| Musculoskeletal Disorders | Exemestane | *ESR1* | rs2813543 | A | Recessive | T-score | Post-menopausal | Oesterreich 2015 | Age, BMI, prior chemotherapy or tamoxifen | Mean Change T score (SD)=VT/VT −1.55 (1.91) vs. WT/WT, WT/VT −0.31 (0.41), p=2.1E−4 | Age, BMI, prior chemotherapy, assessment centre, first five principal components | Main effects model | β coefficient=-0.0057766 | -0.032267 | 0.0207137 | 6.70E-01 |

| | | | | | | | | | | | | | | | | |
|---|---|---|---|---|---|---|---|---|---|---|---|---|---|---|---|---|
| | | | | | | | | | | | | Interaction model | β coefficient=-0.8836432 | -2.33382 | 0.566534 | 2.30E-01 |
| Musculoskeletal Disorders | Letrozole | *ESR1* | rs4870061 | T | Recessive | Bone mineral density | Post-menopausal | Oesterreich 2015 | Age, BMI, prior chemotherapy or tamoxifen | Mean % Change BMD (SD)= VT/VT −10.94 (12.71) vs. WT/WT, WT/VT −3.76 (3.49), p=3.0E−4 | Age, BMI, prior chemotherapy, assessment centre, first five principal components | Main effects model | β coefficient=-0.001866 | -0.0044691 | 0.000737 | 1.60E-01 |
| | | | | | | | | | | | | Interaction model | β coefficient=0.0192075 | -0.0366608 | 0.0750759 | 5.00E-01 |
| Musculoskeletal Disorders | Letrozole | *ESR2* | rs10140457 | C | Additive | Bone mineral density | Post-menopausal | Oesterreich 2015 | Age, BMI, prior chemotherapy or tamoxifen | Mean % Change BMD (SD)= WT/VT 3.08 (6.92) vs. WT/WT −3.43 (3.97), p=3.0E−4 | Age, BMI, prior chemotherapy, assessment centre, first five principal components | Main effects model | β coefficient=-0.0013776 | -0.0049584 | 0.0022032 | 4.50E-01 |
| | | | | | | | | | | | | Interaction model | β coefficient=-0.0603724 | -0.1628594 | 0.0421146 | 2.50E-01 |
| Musculoskeletal Disorders | Anastrozole; Exemestane; Letrozole | *CYP11A1* | rs11632698 | A | Additive | Bone mineral density | Post-menopausal | Rodríguez-Sanz 2015 | Age, BMI, prior chemotherapy or tamoxifen. Patients with scoliosis, on bisphosphonates (BP) treatment, spine scan artifacts and/or those with hip scan artifacts or bilateral prostheses were excluded from the study. | β coefficient (95% CI), P value= 0.94 (0.30–1.58), 0.004 | Age, BMI, prior chemotherapy, assessment centre, first five principal components [those with scoliosis, on bisphosphonates (BP) treatment, spine scan artifacts and/or those with hip scan artifacts or bilateral prostheses were excluded] | Main effects model | β coefficient=0.00063477 | -0.0002871 | 0.0015566 | 1.77E-01 |
| | | | | | | | | | | | | Interaction model | β coefficient=0.00354739 | -0.0074157 | 0.0145105 | 5.26E-01 |

| | | | | | | | | | | | | | | | | |
|---|---|---|---|---|---|---|---|---|---|---|---|---|---|---|---|---|
| Musculoskeletal Disorders | Anastrozole; Exemestane; Letrozole | *CYP11A1* | rs900798 | T | Additive | Bone mineral density | Post-menopausal | Rodríguez-Sanz 2015 | Age, BMI, prior chemotherapy or tamoxifen. Patients with scoliosis, on bisphosphonates (BP) treatment, spine scan artifacts and/or those with hip scan artifacts or bilateral prostheses were excluded from the study. | β coefficient (95% CI), P value= 1.06 (0.39–1.72), 0.002 | Age, BMI, prior chemotherapy, assessment centre, first five principal components [those with scoliosis, on bisphosphonates (BP) treatment, spine scan artifacts and/or those with hip scan artifacts or bilateral prostheses were excluded] | Main effects model | β coefficient=0.00051465 | -0.0004526 | 0.0014819 | 2.97E-01 |
| | | | | | | | | | | | | Interaction model | β coefficient=0.00312909 | -0.008436 | 0.0146942 | 5.96E-01 |
| Musculoskeletal Disorders | Anastrozole; Exemestane; Letrozole | *CYP11A1* | rs4077581 | C | Additive | Bone mineral density | Post-menopausal | Rodríguez-Sanz 2015 | Age, BMI, prior chemotherapy or tamoxifen. Patients with scoliosis, on bisphosphonates (BP) treatment, spine scan artifacts and/or those with hip scan artifacts or bilateral prostheses were excluded from the study. | β coefficient (95% CI), P value= 0.98 (0.30–1.66), 0.005 | Age, BMI, prior chemotherapy, assessment centre, first five principal components [those with scoliosis, on bisphosphonates (BP) treatment, spine scan artifacts and/or those with hip scan artifacts or bilateral prostheses were excluded] | Main effects model | β coefficient=0.00050966 | -0.0004745 | 0.0014938 | 3.10E-01 |
| | | | | | | | | | | | | Interaction model | β coefficient=0.00410373 | -0.0076801 | 0.0158875 | 4.95E-01 |

| | | | | | | | | | | | | | | | | |
|---|---|---|---|---|---|---|---|---|---|---|---|---|---|---|---|---|
| Musculoskeletal Disorders | Anastrozole; Letrozole | *TNFRSF11B* | rs2073618 | C | Recessive | Bone mineral density | Post-menopausal | Wang 2015 | N/R | Mean ± SD, (n) =CC vs. GG: 0.801 ± 0.218, (n = 50) vs. 0.987 ± 0.257 (n = 59), P value=0.049 | Assessment centre, first five principal components | Main effects model | Means ± SD:CC vs. GG. [0.4749674 ±0.1197611 vs.0.4789469 ± 0.1035114], (P=0.6952) Diff in mean=-0.0039795, Std. Err.= 0.0101507, [95% Conf. Interval]= [-0.0239225, 0.0159635], P=0.6952 | | | |
| | | | | | | | | | | | | Interaction model | β coefficient=0.00285258 | -0.0158902 | 0.0215954 | 7.65E-01 |
| Musculoskeletal Disorders | Tamoxifen | *E2F7* | rs310786 | C | Additive | Bone mineral density | Pre- , Peri- & Post-menopausal | Weng 2013 | Age, menopausal status, prior chemotherapy | Log2 [Lumbar bone loss, (min, max)] CC= 24.4 [-77.43, 90.6] vs. CT= 14.76 [-90.42, 92.8] vs. TT= -1.73 [-120, 129.9] (P = 0.043) | Age, menopausal status, prior chemotherapy, assessment centre, first five principal components | Main effects model | β coefficient=0.0007763 | -0.0003125 | 0.0018651 | 1.62E-01 |
| | | | | | | | | | | | | Interaction model | β coefficient=0.00108519 | -0.0135412 | 0.0157116 | 8.84E-01 |
| Musculoskeletal Disorders | Tamoxifen | *POLQ* | rs9862879 | T | Additive | Bone mineral density | Pre- , Peri- & Post-menopausal | Weng 2013 | Age, menopausal status, prior chemotherapy | N/R | Age, menopausal status, prior chemotherapy, assessment centre, first five principal components | Main effects model | β coefficient=-0.00076705 | -0.0019399 | 0.0004058 | 2.00E-01 |
| | | | | | | | | | | | | Interaction model | β coefficient=-0.00064037 | -0.0164943 | 0.0152136 | 9.37E-01 |
| Musculoskeletal Disorders | Tamoxifen | *SLC22A23* | rs4959825 | T | Additive | Bone mineral density | Pre- , Peri- & Post-menopausal | Weng 2013 | Age, menopausal status, prior chemotherapy | N/R | Age, menopausal status, prior chemotherapy, assessment centre, first five principal components | Main effects model | β coefficient=-0.00017619 | -0.0009992 | 0.0006468 | 6.75E-01 |
| | | | | | | | | | | | | Interaction model | β coefficient=0.00470449 | -0.006717 | 0.016126 | 4.19E-01 |

| | | | | | | | | | | | | | | | | |
|---|---|---|---|---|---|---|---|---|---|---|---|---|---|---|---|---|
| Musculoskeletal Disorders | Tamoxifen | *E2F7* | rs310786 | C | Recessive | Bone mineral density | Pre- , Peri- & Post-menopausal | Weng 2013 | Age, menopausal status, prior chemotherapy | Log2 [Lumbar bone loss, (min, max)] CC= 24.4 [-77.43, 90.6] vs. CT= 14.76 [-90.42, 92.8] vs. TT= -1.73 [-120, 129.9] (P = 0.043) | Age, menopausal status, prior chemotherapy, assessment centre, first five principal components | Main effects model | β coefficient=0.00084713 | -0.0028745 | 0.0045687 | 6.55E-01 |
| | | | | | | | | | | | | Interaction model | β coefficient=0.01958579 | -0.0308838 | 0.0700554 | 4.47E-01 |
| Musculoskeletal Disorders | Tamoxifen | *PTCSC2* | rs10983920 | A | Additive | Bone mineral density | Pre- , Peri- & Post-menopausal | Weng 2013 | Age, menopausal status, prior chemotherapy | N/R | Age, menopausal status, prior chemotherapy, assessment centre, first five principal components | Main effects model | β coefficient=-0.00001606 | -0.0011662 | 0.0011341 | 9.78E-01 |
| | | | | | | | | | | | | Interaction model | β coefficient=0.00354694 | -0.0118096 | 0.0189035 | 6.51E-01 |
| Metabolism Disorders | Anastrozole; Exemestane; Letrozole | *ESR1* | XbaI (rs9340799) | A | Recessive | Triglycerides | Post-menopausal | Koukouras 2012 | N/R | Xbal (AA)= 159 ± 42 vs. Xbal (AG+GG)=136 ± 40 [p = 0.044] (mean value ± SD) | Assessment centre, first five principal components | Main effects model | β coefficient=0.000039 | -0.009657 | 0.009735 | 9.94E-01 |
| Metabolism Disorders | Anastrozole; Exemestane; Letrozole | *ESR1* | XbaI (rs9340799) | A | Recessive | Triglycerides | Post-menopausal | Koukouras 2012 | N/R | Xbal (AA)= 159 ± 42 vs. Xbal (AG+GG)=136 ± 40 [p = 0.044] (mean value ± SD) | Assessment centre, first five principal components | Main effects model | Xbal (AA)=1.782029 ± 1.000734 vs. Xbal (AG+GG)= 1.715918 ± .8683153 [p =0.2268] (mean value ± SD), diff in mean=.066111, Std. Err.=.0546713, [95% Conf. Interval]= [-.0411528 .1733748], P=0.2268 | | | |
| | | | | | | | | | | | | Interaction model | β coefficient=0.06775764 | -0.033992 | 0.1695073 | 1.92E-01 |
| Metabolism Disorders | Anastrozole; Exemestane; Letrozole | *ESR1* | PvuII (rs2234693) | C | Recessive | Triglycerides | Post-menopausal | Koukouras 2012 | N/R | Pvull (CC)= 161 ± 47 vs. Pvull (TC+TT)= 133 ± 39 [p = 0.015] (mean value ± SD) | Assessment centre, first five principal components | Main effects model | β coefficient=-0.00064186 | -0.0124033 | 0.0111195 | 9.15E-01 |

| | | | | | | | | | | | | | | | |
|---|---|---|---|---|---|---|---|---|---|---|---|---|---|---|---|
| Metabolism Disorders | Anastrozole; Exemestane; Letrozole | *ESR1* | PvuII (rs2234693) | C | Recessive | Triglycerides | Post-menopausal | Koukouras 2012 | N/R | PvuII (CC)= 161 ± 47 vs. PvuII (TC+TT)= 133 ± 39 [p = 0.015] (mean value ± SD) | Assessment centre, first five principal components | Main effects model | PvuII (CC)= 1.7171 ± .9579613 vs. PvuII (TC+TT)=1.750289 ± .9171709 [p=0.6154] (mean value ± SD), diff in mean=-.033189, Std. Err.=.0660513, [95% Conf. Interval]= [-.1627803 .0964023], P =0.6154 | | | |
| | | | | | | | | | | | | Interaction model | β coefficient=-0.03411345 | -0.1569839 | 0.088757 | 5.86E-01 |
| Metabolism Disorders | Anastrozole; Exemestane; Letrozole | *ESR1* | XbaI (rs9340799) | A | Recessive | LDL | Post-menopausal | Koukouras 2012 | N/R | XbaI (AA)= 157 ± 20 vs. XbaI (AG+GG)= 142 ± 23 [p = 0.025] (mean value ± SD) | Assessment centre, first five principal components | Main effects model | β coefficient=-0.00133949 | -0.0110291 | 0.0083501 | 7.86E-01 |
| Metabolism Disorders | Anastrozole; Exemestane; Letrozole | *ESR1* | XbaI (rs9340799) | A | Recessive | LDL | Post-menopausal | Koukouras 2012 | N/R | XbaI (AA)= 157 ± 20 vs. XbaI (AG+GG)= 142 ± 23 [p = 0.025] (mean value ± SD) | Assessment centre, first five principal components | Main effects model | XbaI (AA)=3.746609 ± .9033583 vs. XbaI (AG+GG)= 3.769003 ± .878145 [p = 0.6700] (mean value ± SD), diff in mean=-.022394, Std. Err.=.0525323, [95% Conf. Interval]= [-.1254613 .0806733], P =0.6700 | | | |
| | | | | | | | | | | | | Interaction model | β coefficient=-0.02080825 | -0.1224952 | 0.0808787 | 6.88E-01 |
| Metabolism Disorders | Anastrozole; Exemestane; Letrozole | *ESR1* | PvuII (rs2234693) | C | Recessive | LDL | Post-menopausal | Koukouras 2012 | N/R | PvuII (CC)= 155 ± 20 vs. PvuII (TC+TT)= 141 ± 24 [p = 0.026] (mean value ± SD) | Assessment centre, first five principal components | Main effects model | β coefficient=-0.00120545 | -0.0129589 | 0.010548 | 8.41E-01 |

| | | | | | | | | | | | | | | | | |
|---|---|---|---|---|---|---|---|---|---|---|---|---|---|---|---|---|
| Metabolism Disorders | Anastrozole; Exemestane; Letrozole | *ESR1* | PvuII (rs2234693) | C | Recessive | LDL | Post-menopausal | Koukouras 2012 | N/R | PvuII (CC)= 155 ± 20 vs. PvuII (TC+TT)= 141 ± 24 [p = 0.026] (mean value ± SD) | Assessment centre, first five principal components | Main effects model | PvuII (CC)= 3.791319 ± .836355 vs. PvuII (TC+TT)= 3.751296 ± .9019727 [p = 0.5286] (mean value ± SD), diff in mean=.040023, Std. Err.= .0634987, [95% Conf. Interval]= [-.0845602 .1646062], P =0.5286 | | | |
| | | | | | | | | | | | | Interaction model | β coefficient=0.04104445 | -0.0818835 | 0.1639724 | 5.13E-01 |
| Metabolism Disorders | Tamoxifen | *ESR1* | XbaI (rs9340799) | G | Recessive | Total cholesterol | Post-menopausal | Ntukidem 2008 | N/R (patients were neither perimenopausal nor were taking concomitant lipid-lowering medications) | [Change in serum lipid particle concentration (mg/dl) with 95% CI, p-values between baseline and 4 months: AA/AG vs the GG alleles. Total cholesterol: AA [−22 (−30, −14)], AG [−20 (−30, −10)], GG [−40 (−58, −22)], a statistically significant difference for the AA/AG versus GG genotypes (P=0.03] | Assessment centre, first five principal components [those taking lipid lowering meds were excluded] | Main effects model | β coefficient=-0.00622678 | -0.0249079 | 0.0124544 | 5.14E-01 |
| | | | | | | | | | | | | Interaction model | β coefficient=-0.03504523 | -0.2813176 | 0.2112272 | 7.80E-01 |

| | | | | | | | | | | | | | | | | |
|---|---|---|---|---|---|---|---|---|---|---|---|---|---|---|---|---|
| Metabolism Disorders | Tamoxifen | *ESR1* | XbaI (rs9340799) | G | Additive | Triglycerides | Pre-menopausal | Ntukidem 2008 | N/R (patients were neither perimenopausal nor were taking concomitant lipid-lowering medications) | [Change in serum lipid particle concentration (mg/dl) with 95% CI, p-values between baseline and 4 months: (−) 7mg/dl, (+) 46mg/dl, and (+) 163 mg/dl for women who carry the AA, AG, and GG genotypes, respectively (P=0.002) | Assessment centre, first five principal components [those taking lipid lowering meds were excluded] | Main effects model | β coefficient=0.0044995 | -0.0047509 | 0.0137499 | 3.40E-01 |
| | | | | | | | | | | | | Interaction model | β coefficient=-0.0383777 | -0.1764855 | 0.0997302 | 5.90E-01 |
| Metabolism Disorders | Tamoxifen | *ESR1* | XbaI (rs9340799) | G | Additive | HDL cholesterol | Pre-menopausal | Ntukidem 2008 | N/R (patients were neither perimenopausal nor were taking concomitant lipid-lowering medications) | [Change in serum lipid particle concentration (mg/dl) with 95% CI, p-values between baseline and 4 months: (+) 5mg/dl, (−) 0.6mg/dl, and (−) 4 mg/dl for the AA, AG, and GG genotypes, respectively (P=0.004 for gene–dose effect). | Assessment centre, first five principal components [those taking lipid lowering meds were excluded] | Main effects model | β coefficient=-0.0031129 | -0.0077855 | 0.0015598 | 1.90E-01 |
| | | | | | | | | | | | | Interaction model | β coefficient=-0.0063933 | -0.0779233 | 0.0651367 | 8.60E-01 |
| Metabolism Disorders | Tamoxifen | *ESR2* | ESR2-02 (rs4986938) | A | Additive | Triglycerides | Pre-menopausal | Ntukidem 2008 | N/R (patients were neither perimenopausal nor were taking concomitant lipid-lowering medications) | [Change in serum lipid particle concentration (mg/dl) with 95% CI, p-values between baseline and 4 months: (+) 97 mg/dl, (+) 37 mg/dl, and (+) 7 mg/dl for the GG, AG, and AA genotypes in ESR2-02, respectively (P=0.04 for gene–dose effect) | Assessment centre, first five principal components [those taking lipid lowering meds were excluded] | Main effects model | β coefficient=0.003462 | -0.0056922 | 0.0126163 | 4.60E-01 |

| | | | | | | | | | | | | | | | | |
|---|---|---|---|---|---|---|---|---|---|---|---|---|---|---|---|---|
| | | | | | | | | | | | | Interaction model | β coefficient=0.0941782 | -0.0509734 | 0.2393298 | 2.00E-01 |
| Metabolism Disorders | Tamoxifen | *ESR2* | ESR2-02 (rs4986938) | A | Additive | Triglycerides | Post-menopausal | Ntukidem 2008 | N/R (patients were neither perimenopausal nor were taking concomitant lipid-lowering medications) | [Change in serum lipid particle concentration (mg/dl) with 95% CI, p-values between baseline and 4 months: (+) 71 mg/dl, (+) 8 mg/dl, and (−) 15 mg/dl from baseline for the GG, AG, and AA genotypes in ESR2-02, respectively (P=0.01 for gene—dose effect) | Assessment centre, first five principal components [those taking lipid lowering meds were excluded] | Main effects model | β coefficient=-0.00215755 | -0.0091482 | 0.0048331 | 5.45E-01 |
| | | | | | | | | | | | | Interaction model | β coefficient=-0.02455295 | -0.1191859 | 0.07008 | 6.11E-01 |
| Metabolism Disorders | Letrozole | *CYP19A1* | rs10046 | A | Recessive | Triglycerides | Post-menopausal | Santa-Maria 2016 | Age, BMI, race, change in plasma estradiol, and prior hormonal therapy and tamoxifen use | Mean Change, mg/dL (SE), P-value= −36.4 (7.8), 0.0000069. Only Caucasian women: -30.92 (8.2), 0.00023. Without adjusting for changes in plasma estradiol concentrations: -29.8 (7.3), 0.000082 | Age, BMI, HRT, assessment centre, first five principal components | Main effects model | β coefficient=-0.0069599 | -0.0171134 | 0.0031936 | 1.80E-01 |
| | | | | | | | | | | | | Interaction model | β coefficient=0.1383645 | -0.0827408 | 0.3594697 | 2.20E-01 |
| Metabolism Disorders | Letrozole | *CYP19A1* | rs2289105 | C | Recessive | Triglycerides | Post-menopausal | Santa-Maria 2016 | Age, BMI, race, change in plasma estradiol, and prior hormonal therapy and tamoxifen use | Mean Change, mg/dL (SE), P-value= −36.45 (7.8), 0.0000074. Only Caucasian women: -30.98 (8.2), 0.00024. Without adjusting for changes in plasma estradiol concentrations: -29.8 (7.4), 0.000085 | Age, BMI, HRT, assessment centre, first five principal components | Main effects model | β coefficient=-0.0072335 | -0.0173706 | 0.0029035 | 1.60E-01 |

| | | | | | | | | | | | | | | | | |
|---|---|---|---|---|---|---|---|---|---|---|---|---|---|---|---|---|
| | | | | | | | | | | | | Interaction model | β coefficient=0.1322193 | -0.0882891 | 0.3527277 | 2.40E-01 |
| Metabolism Disorders | Letrozole | *CYP19A1* | rs3759811 | C | Recessive | Triglycerides | Post-menopausal | Santa-Maria 2016 | Age, BMI, race, change in plasma estradiol, and prior hormonal therapy and tamoxifen use | Mean Change, mg/dL (SE), P-value= −32.3 (9.0), 0.00045. Only Caucasian women: -33 (9.4) 0.00065. Without adjusting for changes in plasma estradiol concentrations: -30.8 (8.5), 0.00043 | Age, BMI, HRT, assessment centre, first five principal components | Main effects model | β coefficient=-0.0076403 | -0.0180249 | 0.0027442 | 1.50E-01 |
| | | | | | | | | | | | | Interaction model | β coefficient=0.1019051 | -0.127324 | 0.3311343 | 3.80E-01 |
| Metabolism Disorders | Letrozole | *CYP19A1* | rs700518 | G | Recessive | Triglycerides | Post-menopausal | Santa-Maria 2016 | Age, BMI, race, change in plasma estradiol, and prior hormonal therapy and tamoxifen use | Mean Change, mg/dL (SE), P-value= −32.3 (8.4), 0.00019 | Age, BMI, HRT, assessment centre, first five principal components | Main effects model | β coefficient=-0.0075438 | -0.0179292 | 0.0028417 | 1.50E-01 |
| | | | | | | | | | | | | Interaction model | β coefficient=0.1018087 | -0.1274206 | 0.3310379 | 3.80E-01 |
| Metabolism Disorders | Letrozole | *CYP19A1* | rs4775936 | C | Recessive | Triglycerides | Post-menopausal | Santa-Maria 2016 | Age, BMI, race, change in plasma estradiol, and prior hormonal therapy and tamoxifen use | Mean Change, mg/dL (SE), P-value= −33.2 (8.9), 0.00028 | Age, BMI, HRT, assessment centre, first five principal components | Main effects model | β coefficient=0.0084503 | -0.0018386 | 0.0187391 | 1.10E-01 |
| | | | | | | | | | | | | Interaction model | β coefficient=-0.0955153 | -0.3611857 | 0.1701552 | 4.80E-01 |
| Metabolism Disorders | Letrozole | *CYP19A1* | rs4646 | A | Additive | HDL cholesterol | Post-menopausal | Santa-Maria 2016 | Age, BMI, race, change in plasma estradiol, and prior hormonal therapy and tamoxifen use | Mean Change, mg/dL (SE), P-value= −4.2 (1.16), 0.00043 | Age, BMI, HRT, assessment centre, first five principal components | Main effects model | β coefficient=-0.0033432 | -0.00667 | -1.65E-05 | 4.90E-02 |
| | | | | | | | | | | | | Interaction model | β coefficient=-0.0321437 | -0.1167246 | 0.0524372 | 4.60E-01 |

| | | | | | | | | | | | | | | | | |
|---|---|---|---|---|---|---|---|---|---|---|---|---|---|---|---|---|
| Metabolism Disorders | Letrozole | *CYP19A1* | rs749292 | A | Recessive | Triglycerides | Post-menopausal | Santa-Maria 2016 | Age, BMI, race, change in plasma estradiol, and prior hormonal therapy and tamoxifen use | Mean Change, mg/dL (SE), P-value= −39.3 (8.6), 0.000012. Only Caucasian women: -37.47 (9.3), 0.000095. Without adjusting for changes in plasma estradiol concentrations: -32.8 (8.3), 0.00012 | Age, BMI, HRT, assessment centre, first five principal components | Main effects model | β coefficient=-0.007234 | -0.0187133 | 0.0042453 | 2.20E-01 |
| | | | | | | | | | | | | Interaction model | β coefficient=0.1593088 | -0.1022963 | 0.4209139 | 2.30E-01 |
| Metabolism Disorders | Letrozole | *CYP19A1* | rs749292 | A | Additive | Triglycerides | Post-menopausal | Santa-Maria 2016 | Age, BMI, race, change in plasma estradiol, and prior hormonal therapy and tamoxifen use | Mean Change, mg/dL (SE), P-value= −20.2 (4.7), 0.000037. Only Caucasian women: -19.51 (5.0), 0.00017. Without adjusting for changes in plasma estradiol concentrations: -17.1 (4.5), 0.00021 | Age, BMI, HRT, assessment centre, first five principal components | Main effects model | β coefficient=-0.005955 | -0.0124772 | 0.0005672 | 7.40E-02 |
| | | | | | | | | | | | | Interaction model | β coefficient=0.092874 | -0.068907 | 0.254655 | 2.60E-01 |
| Metabolism Disorders | Letrlipid [taking letrozole and on lipid-altering medications] | *CYP19A1* | rs749292 | A | Additive | HDL cholesterol | Post-menopausal | Santa-Maria 2016 | Age, BMI, race, change in plasma estradiol, and prior hormonal therapy and tamoxifen use | Mean Change, mg/dL (SE), P-value= 6.7 (1.4), 0.000037. Only Caucasian women: 6 (1.3), 0.000065. | Age, BMI, HRT, assessment centre, first five principal components | Main effects model | β coefficient=0.0016827 | -0.0012467 | 0.0046122 | 2.60E-01 |
| | | | | | | | | | | | | Interaction model | β coefficient=0.0155252 | -0.1687571 | 0.1998074 | 8.70E-01 |
| Metabolism Disorders | Letrlipid [taking letrozole and on lipid-altering medications] | *CYP19A1* | rs1062033 | G | Additive | HDL cholesterol | Post-menopausal | Santa-Maria 2016 | Age, BMI, race, change in plasma estradiol, and prior hormonal therapy and tamoxifen use | Mean Change, mg/dL (SE), P-value= 6.9 (1.5), 0.000043. Only Caucasian women: 6.2 (1.3), 0.000023 | Age, BMI, HRT, assessment centre, first five principal components | Main effects model | β coefficient=0.0023691 | -0.0005492 | 0.0052873 | 1.10E-01 |
| | | | | | | | | | | | | Interaction model | β coefficient=0.0132003 | -0.1710833 | 0.1974839 | 8.90E-01 |

| | | | | | | | | | | | | | | | | |
|---|---|---|---|---|---|---|---|---|---|---|---|---|---|---|---|---|
| Metabolism Disorders | Letrlipid [taking letrozole and on lipid-altering medications] | *CYP19A1* | rs1062033 | G | Dominant | HDL cholesterol | Post-menopausal | Santa-Maria 2016 | Age, BMI, race, change in plasma estradiol, and prior hormonal therapy and tamoxifen use | Mean Change, mg/dL (SE), P-value= Only Caucasian women: 8.26, 0.00035 | Age, BMI, HRT, assessment centre, first five principal components | Main effects model | β coefficient=0.0022315 | -0.0022983 | 0.0067614 | 3.34E-01 |
| | | | | | | | | | | | | Interaction model | β coefficient=0.0257468 | -0.2146254 | 0.266119 | 8.30E-01 |
| Metabolism Disorders | Letrlipid [taking letrozole and on lipid-altering medications] | *CYP19A1* | rs1008805 | G | Additive | HDL cholesterol | Post-menopausal | Santa-Maria 2016 | Age, BMI, race, change in plasma estradiol, and prior hormonal therapy and tamoxifen use | Mean Change, mg/dL (SE), P-value= Only Caucasian women: -5.01 (1.6), 0.003. Without adjusting for changes in plasma estradiol concentrations: -5.8 (1.6), 0.00054 | Age, BMI, HRT, assessment centre, first five principal components | Main effects model | β coefficient=-0.0029017 | -0.0058462 | 0.0000427 | 5.30E-02 |
| | | | | | | | | | | | | Interaction model | β coefficient=0.0410825 | -0.1435086 | 0.2256737 | 6.60E-01 |
| Metabolism Disorders | Letrlipid [taking letrozole and on lipid-altering medications] | *CYP19A1* | rs1008805 | G | Dominant | HDL cholesterol | Post-menopausal | Santa-Maria 2016 | Age, BMI, race, change in plasma estradiol, and prior hormonal therapy and tamoxifen use | Mean Change, mg/dL (SE), P-value= −6.6 (1.7), 0.00037. Only Caucasian women: -6.96 (2.2), 0.003 | Age, BMI, HRT, assessment centre, first five principal components | Main effects model | β coefficient=-0.0048915 | -0.0092489 | -0.000534 | 2.80E-02 |
| | | | | | | | | | | | | Interaction model | β coefficient=-0.0003716 | -0.2550321 | 0.2542889 | 1.00E+00 |
| Metabolism Disorders | Letrlipid [taking letrozole and on lipid-altering medications] | *CYP19A1* | rs749292 | A | Dominant | HDL cholesterol | Post-menopausal | Santa-Maria 2016 | Age, BMI, race, change in plasma estradiol, and prior hormonal therapy and tamoxifen use | Mean Change, mg/dL (SE), P-value= 9.5 (2.4), 0.0003. Only Caucasian women: 8.26 (2.1), 0.00035 | Age, BMI, HRT, assessment centre, first five principal components | Main effects model | β coefficient=0.0017033 | -0.0027562 | 0.0061628 | 4.50E-01 |
| | | | | | | | | | | | | Interaction model | β coefficient=0.0290638 | -0.2113052 | 0.2694327 | 8.10E-01 |
| Metabolism Disorders | Letrlipid [taking letrozole and on lipid-altering medications] | *CYP19A1* | rs10046 | C | Additive | HDL cholesterol | Post-menopausal | Santa-Maria 2016 | Age, BMI, race, change in plasma estradiol, and prior hormonal therapy and tamoxifen use | Mean Change, mg/dL (SE), P-value= −6.2 (1.6), 0.00046 | Age, BMI, HRT, assessment centre, first five principal components | Main effects model | β coefficient=-0.0027602 | -0.0056729 | 0.0001526 | 6.30E-02 |
| | | | | | | | | | | | | Interaction model | β coefficient=-0.0786367 | -0.2607772 | 0.1035037 | 4.00E-01 |

| | | | | | | | | | | | | | | | | |
|---|---|---|---|---|---|---|---|---|---|---|---|---|---|---|---|---|
| Metabolism Disorders | Letrlipid [taking letrozole and on lipid-altering medications] | *CYP19A1* | rs2289105 | T | Additive | HDL cholesterol | Post-menopausal | Santa-Maria 2016 | Age, BMI, race, change in plasma estradiol, and prior hormonal therapy and tamoxifen use | Mean Change, mg/dL (SE), P-value= −6.2 (1.6), 0.00046 | Age, BMI, HRT, assessment centre, first five principal components | Main effects model | β coefficient=-0.0028412 | -0.0057554 | 0.000073 | 5.60E-02 |
| | | | | | | | | | | | | Interaction model | β coefficient=-0.0785562 | -0.2606965 | 0.1035841 | 4.00E-01 |

Supplementary Table S6

| System Organ Class | Regimen or Drug(s) | Gene | SNP rs# ID | Genotype | Statistical genetic model | Toxicity endpoint or Parameter | Menopausal status | Original study | | | Replication attempt in UK Biobank | | | | | |
|---|---|---|---|---|---|---|---|---|---|---|---|---|---|---|---|---|
| | | | | | | | | Original study | Adjustement | Effect size [OR or HR (IC95%, UC95%), p value] | Adjustment | Statistical model | OR | IC95% | UC95% | p_value |
| **Musculoskeletal Disorders** | **Anastrozole; Exemestane; Letrozole** | **CYP19A1** | **rs10046** | **A** | **Recessive** | **>5% bone loss (T-Score <= – o.5 )** | **Post-menopausal** | **Baatjes 2020** | **N/R** | **OR=7.37 (1.101-49.336), p=0.04** | **Assessment centre, first five principal components** | **Main effects model** | **0.9493356** | **0.9253461** | **0.97395** | **6.80E-05** |
| | | | | | | | | | | | | Interaction model | 0.9675772 | 0.7322109 | 1.2786 | 8.20E-01 |
| Musculoskeletal Disorders | Tamoxifen | CYP19A1 | rs4646 | A | Additive | Fractures & Osteoporosis | Post-menopausal | Leyland-Jones 2015 [1] | Age, BMI, smoking, HRT, bisphosphonates use, personal history of osteoporosis or bone fractures | HR= 0.80 (0.66, 0.99) | Age, BMI, smoking, HRT, Bisphosphonates, personal history of osteoporosis or bone fractures, assessment centre, first five principal | Main effects model | 1.04010448 | 0.9653562 | 1.12064 | 3.01E-01 |
| | | | | | | | | | | | | Interaction model | 2.75883337 | 0.9777463 | 7.78439 | 5.52E-02 |
| Musculoskeletal Disorders | Tamoxifen | CYP19A1 | rs4646 | A | Dominant | Fractures & Osteoporosis | Post-menopausal | Leyland-Jones 2015 [1] | Age, BMI, smoking, HRT, bisphosphonates use, personal history of osteoporosis or bone fractures | HR=0.76 (0.59, 0.98) | Age, BMI, smoking, HRT, Bisphosphonates, personal history of osteoporosis or bone fractures, assessment centre, first five principal | Main effects model | 1.00074542 | 0.911755 | 1.09842 | 9.87E-01 |
| | | | | | | | | | | | | Interaction model | 3.46356567 | 0.6594666 | 18.1909 | 1.42E-01 |
| Musculoskeletal Disorders | Tamoxifen | CYP19A1 | rs10046 | C | Dominant | Fractures & Osteoporosis | Post-menopausal | Leyland-Jones 2015 [1] | Age, BMI, smoking, HRT, bisphosphonates use, personal history of osteoporosis or bone fractures | HR=1.42 (1.05, 1.93) | Age, BMI, smoking, HRT, Bisphosphonates, personal history of osteoporosis or bone fractures, assessment centre, first five principal | Main effects model | 1.01784256 | 0.9180846 | 1.12844 | 7.37E-01 |
| | | | | | | | | | | | | Interaction model | 1.20140861 | 0.2276916 | 6.3392 | 8.29E-01 |
| Musculoskeletal Disorders | Tamoxifen | CYP19A1 | rs10046 | C | Additive | Fractures & Osteoporosis | Post-menopausal | Leyland-Jones 2015 [1] | Age, BMI, smoking, HRT, bisphosphonates use, personal history of osteoporosis or bone fractures | HR=1.28 (1.07, 1.52) | Age, BMI, smoking, HRT, Bisphosphonates, personal history of osteoporosis or bone fractures, assessment centre, first five principal | Main effects model | 1.0462356 | 0.9798895 | 1.11707 | 1.76E-01 |
| | | | | | | | | | | | | Interaction model | 1.17220927 | 0.4223221 | 3.25362 | 7.60E-01 |
| Musculoskeletal Disorders | Letrozole | CYP19A1 | rs936308 | G | Dominant | Fractures & Osteoporosis | Post-menopausal | Leyland-Jones 2015 [1] | Age, BMI, smoking, HRT, bisphosphonates use, personal history of osteoporosis or bone fractures | HR=0.73 (0.54, 0.99) | Age, BMI, smoking, HRT, Bisphosphonates, personal history of osteoporosis or bone fractures, assessment centre, first five principal | Main effects model | 1.06042402 | 0.9561191 | 1.17611 | 2.67E-01 |
| | | | | | | | | | | | | Interaction model | 0.4908334 | 0.1064576 | 2.26304 | 3.60E-01 |
| Musculoskeletal Disorders | Letrozole | ESR2 | rs4986938 | A | Dominant | Fractures & Osteoporosis | Post-menopausal | Leyland-Jones 2015 (2) | Age, BMI, smoking, HRT, bisphosphonates use, personal history of osteoporosis or bone fractures | HR=1.37 (1.01,1.84) | Age, BMI, smoking, HRT, Bisphosphonates, personal history of osteoporosis or bone fractures, assessment centre, first five principal | Main effects model | 0.97296801 | 0.8850735 | 1.06959 | 5.71E-01 |
| | | | | | | | | | | | | Interaction model | 1.069043 | 0.3647232 | 3.13348 | 9.00E-01 |

| | | | | | | | | | | | | | | | |
|---|---|---|---|---|---|---|---|---|---|---|---|---|---|---|---|
| Musculoskeletal Disorders | Letrozole | ESR1 | rs2077647 | C | Additive | Fractures & Osteoporosis | Post-menopausal | Leyland-Jones 2015 (2) | Age, BMI, smoking, HRT, bisphosphonates use, personal history of osteoporosis or bone fractures | HR=0.82 (0.68,0.99) | Age, BMI, smoking, HRT, Bisphosphonates, personal history of osteoporosis or bone fractures, assessment centre, first five principal | Main effects model | 0.97922223 | 0.9170176 | 1.04565 | 5.31E-01 |
| | | | | | | | | | | | | Interaction model | 1.49561917 | 0.3785692 | 5.90876 | 5.66E-01 |
| Musculoskeletal Disorders | Letrozole | ESR1 | rs2077647 | C | Dominant | Fractures & Osteoporosis | Post-menopausal | Leyland-Jones 2015 (2) | Age, BMI, smoking, HRT, bisphosphonates use, personal history of osteoporosis or bone fractures | HR=0.75 (0.58,0.98) | Age, BMI, smoking, HRT, Bisphosphonates, personal history of osteoporosis or bone fractures, assessment centre, first five principal | Main effects model | 1.00131423 | 0.9020355 | 1.11152 | 9.80E-01 |
| | | | | | | | | | | | | Interaction model | 1.10078559 | 0.1099067 | 11.0251 | 9.35E-01 |
| Musculoskeletal Disorders | Anastrozole; Letrozole | CYP19A1 | rs4646 | G | Recessive | Osteoporosis | Post-menopausal | Mazzuca 2016 | Age | OR=36.63 (1.99-673.8), P = 0.0154 [Total sample n=11 (24.4)/(n=45), GG n=11 (45.8)/(n=24), GT n=0 (0.0)/(n=19), TT n=0 (0.0)/(n=2) | Age, assessment centre, first five principal components [those who are positive for osteoporotic, before the therapy, being excluded] | Main effects model | 0.98468453 | 0.9093068 | 1.06631 | 7.04E-01 |
| | | | | | | | | | | | | Interaction model | 0.77122364 | 0.3388614 | 1.75525 | 5.36E-01 |
| Musculoskeletal Disorders | Anastrozole; Letrozole | TNFRSF11B | rs2073618 | C | Recessive | Osteoporosis [<−2.5] | Post-menopausal | Wang 2015 | N/R | OR=2.76 (1.13, 6.73), P = 0.0259 [CC n (%) vs. GG n (%), p-value: 18 (36.0)/(n=50) vs. 10(16.9)/(n=59), | Assessment centre, first five principal components | Main effects model | 1.08332729 | 0.983686 | 1.19306 | 1.04E-01 |
| | | | | | | | | | | | | Interaction model | 0.62974768 | 0.2417269 | 1.64062 | 3.44E-01 |
| Musculoskeletal Disorders | Anastrozole; Letrozole | TNFSF11 | rs7984870 | C | Recessive | Osteoporosis [<−2.5] | Post-menopausal | Wang 2015 | N/R | OR=4.19 (1.16, 15.17), P = 0.0288 [CC n (%) vs GG n (%), p-value = 23 (30.2)/(n=76) vs. 3 (9.4)/(n=32), | Assessment centre, first five principal components | Main effects model | 0.96696838 | 0.876096 | 1.06727 | 5.05E-01 |
| | | | | | | | | | | | | Interaction model | 0.77266041 | 0.3176162 | 1.87964 | 5.70E-01 |
| **Musculoskeletal Disorders** | **Anastrozole; Letrozole** | **TNFRSF11B** | **rs2073618** | **C** | **Recessive** | **Osteopenia [−2.5 –−1.0]** | **Post-menopausal** | **Wang 2015** | **N/R** | **OR=2.61 (1.12-6.11), P = 0.0268 [CC n (%) vs. GG n (%), p-value = 20 (40.0)/(n=50) vs. 12 (20.3)/(n=59), 0.034]** | **Assessment centre, first five principal components** | **Main effects model** | **1.073256** | **1.043746** | **1.1036** | **6.70E-07** |
| | | | | | | | | | | | | Interaction model | 1.17818 | 0.856125 | 1.62139 | 3.10E-01 |
| Musculoskeletal Disorders | Anastrozole; Letrozole | TNFSF11 | rs7984870 | C | Recessive | Osteopenia [−2.5 –−1.0] | Post-menopausal | Wang 2015 | N/R | OR=3.15 (1.16, 8.55), P = 0.0241 [CC n (%) vs. GG n (%), p-value = 32 (42.1)/(n=76) vs. 6 (18.8)/(n=32), | Assessment centre, first five principal components | Main effects model | 1.013846 | 0.9860768 | 1.0424 | 3.30E-01 |
| | | | | | | | | | | | | Interaction model | 0.9206356 | 0.6726205 | 1.2601 | 6.10E-01 |
| **Musculoskeletal Disorders** | **Anastrozole; Letrozole** | **TNFRSF11B** | **rs2073618** | **C** | **Recessive** | **T-score [>-1.0 SD (Normal)]** | **Post-menopausal** | **Wang 2015** | **N/R** | **OR=0.1878 (0.0813, 0.4334), P = 0.0001 [CC n (%) vs. GG n (%), p-value = 12 (24.0)/(n=50) vs. 37 (62.7)/(n=59), 9.20E-5]NOT 0.38 (0.23; 0.65)** | **Assessment centre, first five principal components** | **Main effects model** | **0.9270607** | **0.9018328** | **0.95299** | **7.40E-08** |

| | | | | | | | | | | | | | | | |
|---|---|---|---|---|---|---|---|---|---|---|---|---|---|---|---|
| | | | | | | | | | | | | Interaction model | 0.9004484 | 0.6552269 | 1.23745 | 5.20E-01 |
| Musculoskeletal Disorders | Anastrozole; Letrozole | TNFSF11 | rs7984870 | C | Recessive | T-score [>-1.0 SD (Normal)] | Post-menopausal | Wang 2015 | N/R | OR=0.1494 (0.0595 to 0.3750), P = 0.0001 [CC n (%) vs. GG n (%), p-value = 21 (27.2)/(n=76) vs. 23 (71.8)/(n=32), 3.15E -5] NOT 0.38 (0.25–0.59) | Assessment centre, first five principal components | Main effects model | 0.9891221 | 0.96231 | 1.01668 | 4.40E-01 |
| | | | | | | | | | | | | Interaction model | 1.123564 | 0.8234567 | 1.53305 | 4.60E-01 |
| Musculoskeletal Disorders | Anastrozole; Letrozole | TNFSF11 | rs7984870 | C | Additive | Musculoskeletal Adverse Events | Post-menopausal | Wang 2015 | N/R | OR=1.787 (1.359–2.351), p=3.07E-5 | Assessment centre, first five principal components | Main effects model | 1.01350595 | 0.7835577 | 1.31094 | 9.19E-01 |
| | | | | | | | | | | | | Interaction model | 1.25497007 | 0.0738986 | 21.3123 | 8.75E-01 |
| Musculoskeletal Disorders | Anastrozole; Letrozole | TNFSF11 | rs7984870 | C | Recessive | Musculoskeletal Adverse Events | Post-menopausal | Wang 2015 | N/R | OR=3.259 (1.843–5.763), p=2.19E-4 | Assessment centre, first five principal components | Main effects model | 0.84278578 | 0.5254949 | 1.35166 | 4.78E-01 |
| | | | | | | | | | | | | Interaction model | 1 (omitted) | | | |
| Musculoskeletal Disorders | Anastrozole; Letrozole | TNFRSF11B | rs2073618 | C | Additive | Musculoskeletal Adverse Events | Post-menopausal | Wang 2015 | N/R | OR=1.623 (1.233–2.141), p=6.09E-4 | Assessment centre, first five principal components | Main effects model | 0.9461669 | 0.7303715 | 1.22572 | 6.75E-01 |
| | | | | | | | | | | | | Interaction model | 1.42743124 | 0.079961 | 25.4819 | 8.09E-01 |
| Musculoskeletal Disorders | Anastrozole; Letrozole | TNFRSF11B | rs2073618 | C | Recessive | Musculoskeletal Adverse Events | Post-menopausal | Wang 2015 | N/R | OR=2.931 (1.624–5.288), p=7.95E-4 | Assessment centre, first five principal components | Main effects model | 0.72068008 | 0.43605 | 1.1911 | 2.01E-01 |
| | | | | | | | | | | | | Interaction model | 1 (omitted) | | | |
| Musculoskeletal Disorders | Anastrozole; Letrozole | ESR1 | PvuII (rs2234693) | T | Additive | Musculoskeletal Adverse Events | Post-menopausal | Wang 2013 | Age, type of third generation AIs, BMI and prior tamoxifen use | OR=0.510 (0.389–0.669), p=9.49E-07 | Age, BMI, assessment centre, first five principal components | Main effects model | 0.93662941 | 0.7243718 | 1.21108 | 6.18E-01 |
| | | | | | | | | | | | | Interaction model | convergence not achieved | | | |
| Musculoskeletal Disorders | Anastrozole; Letrozole | ESR1 | PvuII (rs2234693) | T | Recessive | Musculoskeletal Adverse Events | Post-menopausal | Wang 2013 | Age, type of third generation AIs, BMI and prior tamoxifen use | OR=0.282 (0.147–0.540), p=0.001 | Age, BMI, assessment centre, first five principal components | Main effects model | 0.90766325 | 0.6043561 | 1.36319 | 6.41E-01 |
| | | | | | | | | | | | | Interaction model | convergence not achieved | | | |
| Musculoskeletal Disorders | Anastrozole; Letrozole | ESR1 | XbaI (rs9340799) | G | Additive | Musculoskeletal Adverse Events | Post-menopausal | Wang 2013 | Age, type of third generation AIs, BMI and prior tamoxifen use | OR=0.553 (0.399–0.764), p=3.09E-04 | Age, BMI, assessment centre, first five principal components | Main effects model | 0.92033887 | 0.7012185 | 1.20793 | 5.50E-01 |
| | | | | | | | | | | | | Interaction model | 1 (omitted) | | | |
| Musculoskeletal Disorders | Anastrozole; Letrozole | ESR1 | XbaI (rs9340799) | G | Recessive | Musculoskeletal Adverse Events | Post-menopausal | Wang 2013 | Age, type of third generation AIs, BMI and prior tamoxifen use | OR=0.593 (0.441–0.797) | Age, BMI, assessment centre, first five principal components | Main effects model | 0.96064996 | 0.5495118 | 1.6794 | 8.88E-01 |
| | | | | | | | | | | | | Interaction model | 1 (omitted) | | | |
| Hepatobiliary Disorders | Tamoxifen | CYP17A1 | CYP17 A1 allele (rs743572) | A | Additive | Hepatic steatosis | Pre- , Peri- & Post-menopausal | Ohnishi 2005 | N/R | OR=0.53 (0.33-0.82), p=0.005 | Assessment centre, first five principal components | Main effects model | 0.99484593 | 0.9446191 | 1.04774 | 8.45E-01 |
| | | | | | | | | | | | | Interaction model | 1.1687376 | 0.6481178 | 2.10756 | 6.04E-01 |
| Hepatobiliary Disorders | Tamoxifen | CYP17A1 | CYP17 A2 allele (rs743572) | G | Additive | Hepatic steatosis | Pre- , Peri- & Post-menopausal | Ohnishi 2005 | N/R | OR=1.90 (1.21-2.99) | Assessment centre, first five principal components | Main effects model | 1.00518077 | 0.9544321 | 1.05863 | 8.45E-01 |
| | | | | | | | | | | | | Interaction model | 0.85562405 | 0.4744822 | 1.54293 | 6.04E-01 |
| Hepatobiliary Disorders | Tamoxifen | CYP17A1 | CYP17 A2/A2 genotype (rs743572) | G | Recessive | Hepatic steatosis | Pre- , Peri- & Post-menopausal | Ohnishi 2005 | N/R | OR=3.60 (1.42–9.10) | Assessment centre, first five principal components | Main effects model | 0.99479172 | 0.8985591 | 1.10133 | 9.20E-01 |
| | | | | | | | | | | | | Interaction model | 0.24444076 | 0.0327627 | 1.82376 | 1.69E-01 |

| | | | | | | | | | | | | | | | | |
|---|---|---|---|---|---|---|---|---|---|---|---|---|---|---|---|---|
| Hepatobiliary Disorders | Tamoxifen | CYP2D6 | CYP2D6*41 [combination of 2988G>A (rs28371725), 2850C>T (rs16947), –1584C (rs1080985)] | A, T, C | Additive | Hepatic steatosis | Pre- , Peri- & Post-menopausal | Wickramage 2017 | N/R | (P=0.029) | Assessment centre, first five principal components | Main effects model | 1 (omitted) | | | |
| | | | | | | | | | | | | Interaction model | 1 (omitted) | | | |
| Vascular Disorders | Tamoxifen | F5 | FV Leiden (rs6025) | A | Dominant | Thromboembolic events | Pre- , Peri- & Post-menopausal | Garber 2010 | Smoking, personal history and family history of thromboembolic events | OR=4.73 (2.10, 10.68), p < .001 | Assessment centre, first five principal components, smoking, family history of stroke | Main effects model | 1.617275 | 1.426763 | 1.83323 | 5.60E-14 |
| | | | | | | | | | | | | Interaction model | 1.954492 | 0.7824797 | 4.88197 | 1.50E-01 |
| Vascular Disorders | Tamoxifen | F5 | FV Leiden (rs6025) | A | Dominant | Venous thromboembolism [DVT/PE] | Pre- , Peri- & Post-menopausal | Kovac 2015 | N/R | OR=3.32 (1.18, 9.35), P = 0.023 [VTE and FV Leiden mutation frequency (10/50 vs 7/100; p=0.020)] | Assessment centre, first five principal components | Main effects model | 1.403962 | 1.184609 | 1.66393 | 9.10E-05 |
| | | | | | | | | | | | | Interaction model | 3.017497 | 1.092581 | 8.33375 | 3.30E-02 |
| Vascular Disorders | Tamoxifen | F5, F2 | FV Leiden (rs6025) or FII G 20210A (rs1799963) | A, A | Dominant | Venous thromboembolism [DVT/PE] | Pre- , Peri- & Post-menopausal | Kovac 2015 | N/R | OR=3.50 (1.42, 8.60), P = 0.0063 [Prothrombotic mutations and VTE (14/50 vs 10/100; p=0.009)] | Assessment centre, first five principal components | Main effects model | 1.54 | 1.35 | 1.77 | 4.10E-10 |
| | | | | | | | | | | | | Interaction model | 2.88 | 1.18 | 7.03 | 2.0e-02 |
| Vascular Disorders | Tamoxifen | ESR1 | XbaI (rs9340799) | G | Dominant | Venous thromboembolism [DVT/PE] | Pre- , Peri- & Post-menopausal | Onitilo 2009 | Unadjusted analysis but the association persisted after adjusting for classical risk factors including age at diagnosis and body mass index at enrollment | HR=3.47 (0.97–12.44), p=0.035 | Assessment centre, first five principal components | Main effects model | 1.00775721 | 0.9283208 | 1.09399 | 8.54E-01 |
| | | | | | | | | | | | | Interaction model | 1.10882431 | 0.1372882 | 8.95555 | 9.23E-01 |
| Reproductive System | Tamoxifen | CYP3A4 | CYP3A4*1B (rs2740574) | C | Additive | Endometrial cancer | Pre- , Peri- & Post-menopausal | Chu 2007 | N/R | OR=2.8 (1.4, 5.8), p=0.004 | Assessment centre, first five principal components | Main effects model | 0.87133401 | 0.6779862 | 1.11982 | 2.82E-01 |
| | | | | | | | | | | | | Interaction model | 1 (omitted) | | | |
| Reproductive System | Tamoxifen | ABCB1 | rs1045642 | T | Additive | Gynaecological AEs [endometrial hyperplasia or cancer] | Pre- , Peri- & Post-menopausal | Argalacsova 2017 | N/R | HR=1.0588, p=0.0221: v/v, n/total=19/51, wt/v, n/total=21/127, Wt/wt. | Assessment centre, first five principal components | Main effects model | 1.05371379 | 0.9816733 | 1.13104 | 1.48E-01 |
| | | | | | | | | | | | | Interaction model | 0.91455654 | 0.5804361 | 1.44101 | 7.00E-01 |

| | | | | | | | | | | | | | | | | |
|---|---|---|---|---|---|---|---|---|---|---|---|---|---|---|---|---|
| Reproductive System | Tamoxifen | CYP3A5 | CYP3A5*3 (rs776746) | G | Dominant | Endometrial hyperplasia | Pre- , Peri- & Post-menopausal | Miranda 2021 | BMI, blood group, smoking habit, socioeconomic status, treatment with oral contraceptives, hormone replacement therapies, age at menarche, menopausal status, family history of cancer, age at diagnosis, cancer stage | OR=0.067 (0.009–0.524), p=0.010 | Age, BMI, smoking, HRT, menopausal status, age at menarche, family history of cancer, oral contraceptives, assessment centre, first five principal components | Main effects model | 0.66590009 | 0.2130451 | 2.08136 | 4.84E-01 |
| | | | | | | | | | | | | Interaction model | 101334.3 | 0 | . | 9.90E-01 |
| Reproductive System | Tamoxifen | CYP3A5 | CYP3A5*3 (rs776746) | G | Recessive | Endometrial hyperplasia | Pre- , Peri- & Post-menopausal | Miranda 2021 | BMI, blood group, smoking habit, socioeconomic status, treatment with oral contraceptives, hormone replacement therapies, age at menarche, menopausal status, family history of cancer, age at diagnosis, cancer stage | OR=0.208 (0.047–0.758), p=0.007 | Age, BMI, smoking, HRT, menopausal status, age at menarche, family history of cancer, oral contraceptives, assessment centre, first five principal components | Main effects model | 0.93744399 | 0.7148078 | 1.22942 | 6.41E-01 |
| | | | | | | | | | | | | Interaction model | 1656736 | 0 | . | 9.90E-01 |
| Reproductive System | Tamoxifen | CYP2D6 | CYP2D6*4 (rs1800716) | A | Additive | Endometrial hyperplasia | Post-menopausal | Dieudonné 2014 | N/R | HR=1.88 (1.28–2.77), p=0.0022 | Assessment centre, first five principal components | Main effects model | 1.17170031 | 0.9540495 | 1.439 | 1.31E-01 |
| | | | | | | | | | | | | Interaction model | 1.15565318 | 0.3068843 | 4.35191 | 8.31E-01 |
| Reproductive System | Tamoxifen | CYP2D6 | CYP2D6*4 (rs1800716) | A | Recessive | Endometrial hyperplasia | Post-menopausal | Dieudonné 2014 | N/R | HR=2.786 (0.99–7.86), p=0.0022 | Assessment centre, first five principal components | Main effects model | 0.96108166 | 0.5247483 | 1.76023 | 8.98E-01 |
| | | | | | | | | | | | | Interaction model | 5.53506273 | 0.57215 | 53.547 | 1.39E-01 |
| Reproductive System | Anastrozole; Exemestane; Letrozole | ESR1 | Xbal (rs9340799) | A | Recessive | Endometrial hyperplasia | Post-menopausal | Koukouras 2012 | N/R | Mean ± SD: Xbal (AA)= –4.27 ± 3.3 vs. Xbal (AG+GG)=−1.73 ± 2.2 [p=0.005] | Assessment centre, first five principal components | Main effects model | 0.81085152 | 0.6267122 | 1.04909 | 1.11E-01 |
| | | | | | | | | | | | | Interaction model | 1 (omitted) | | | |
| Psychiatric Disorders | Tamoxifen | UGT2B7 | UGT2B7*2 (rs7439366) | T | Additive | Depression | Pre- , Peri- & Post-menopausal | Al-Mamun 2017 | Age, BMI | OR=2.61, p=0.033 | Age, BMI, assessment centre, first five principal | Main effects model | 0.99783881 | 0.9662089 | 1.0305 | 8.95E-01 |
| | | | | | | | | | | | | Interaction model | 0.92603434 | 0.6140076 | 1.39663 | 7.14E-01 |
| Psychiatric Disorders | Tamoxifen | UGT2B7 | UGT2B7*2 (rs7439366) | T | Additive | Depression | Pre- , Peri- & Post-menopausal | Al-Mamun 2017 | Unadjusted | OR=2.5666 (1.0740- 6.1337) | Assessment centre, first five principal components | Main effects model | 0.99997132 | 0.9684329 | 1.03254 | 9.99E-01 |
| | | | | | | | | | | | | Interaction model | 0.90597373 | 0.6003133 | 1.36727 | 6.38E-01 |
| Psychiatric Disorders | Tamoxifen | CYP2D6 | CYP2D6*4 (rs1800716) | A | Additive | Depression | Pre- , Peri- & Post-menopausal | Al-Mamun 2017 | Age, BMI | OR=0.078, p=0.001 | Age, BMI, assessment centre, first five principal | Main effects model | 1.01626315 | 0.9773365 | 1.05674 | 4.18E-01 |
| | | | | | | | | | | | | Interaction model | 1.1550808 | 0.7115666 | 1.87503 | 5.60E-01 |
| Psychiatric Disorders | Tamoxifen | CYP2D6 | CYP2D6*4 (rs1800716) | A | Additive | Depression | Pre- , Peri- & Post-menopausal | Al-Mamun 2017 | Unadjusted | OR=0.0835 (0.0197- 0.3551) | Assessment centre, first five principal components | Main effects model | 1.01367287 | 0.9750603 | 1.05381 | 4.93E-01 |
| | | | | | | | | | | | | Interaction model | 1.17033815 | 0.7196254 | 1.90334 | 5.26E-01 |
| Psychiatric Disorders | Tamoxifen | CYP2D6 | CYP2D6*4 (rs1800716) | A | Dominant | Depression | Pre- , Peri- & Post-menopausal | Al-Mamun 2017 | Age, BMI | OR=0.074, p=0.0006 | Age, BMI, assessment centre, first five principal | Main effects model | 1.02196733 | 0.9750678 | 1.07112 | 3.65E-01 |
| | | | | | | | | | | | | Interaction model | 1.27080541 | 0.7010576 | 2.30359 | 4.30E-01 |

| | | | | | | | | | | | | | | | |
|---|---|---|---|---|---|---|---|---|---|---|---|---|---|---|---|
| Psychiatric Disorders | Tamoxifen | CYP2D6 | CYP2D6*4 (rs1800716) | A | Dominant | Depression | Pre- , Peri- & Post-menopausal | Al-Mamun 2017 | Unadjusted | OR=0.0789 (0.0186, 0.3353) | Assessment centre, first five principal components | Main effects model | 1.01862956 | 0.9721575 | 1.06732 | 4.38E-01 |
| | | | | | | | | | | | | Interaction model | 1.27482839 | 0.7040252 | 2.30842 | 4.23E-01 |
| Psychiatric Disorders | Tamoxifen | CYP2D6 | CYP2D6*10 (rs1065852) | T | Additive | Depression | Pre- , Peri- & Post-menopausal | Al-Mamun 2017 | Age, BMI | OR=1.77E+08, p=0.0223 | Age, BMI, assessment centre, first five principal | Main effects model | 1.01567489 | 0.9782344 | 1.05455 | 4.17E-01 |
| | | | | | | | | | | | | Interaction model | 1.247782 | 0.7887496 | 1.97396 | 3.40E-01 |
| Psychiatric Disorders | Tamoxifen | CYP2D6 | CYP2D6*10 (rs1065852) | T | Additive | Depression | Pre- , Peri- & Post-menopausal | Al-Mamun 2017 | Unadjusted | OR=27.9818 (1.6064, 487.4241) | Assessment centre, first five principal components | Main effects model | 1.01272692 | 0.9755943 | 1.05127 | 5.07E-01 |
| | | | | | | | | | | | | Interaction model | 1.2701942 | 0.8022887 | 2.01099 | 3.08E-01 |
| Psychiatric Disorders | Tamoxifen | CYP2D6 | CYP2D6*10 (rs1065852) | T | Recessive | Depression | Pre- , Peri- & Post-menopausal | Al-Mamun 2017 | Age, BMI | OR=2.32E+08, p=0.0117 | Age, BMI, assessment centre, first five principal | Main effects model | 1.00886723 | 0.9156496 | 1.11157 | 8.58E-01 |
| | | | | | | | | | | | | Interaction model | 1.081103 | 0.3235373 | 3.61252 | 9.00E-01 |
| Psychiatric Disorders | Tamoxifen | CYP2D6 | CYP2D6*10 (rs1065852) | T | Recessive | Depression | Pre- , Peri- & Post-menopausal | Al-Mamun 2017 | Unadjusted | OR=37.2020 (2.2369-618.7153) | Assessment centre, first five principal components | Main effects model | 1.00504463 | 0.9126214 | 1.10683 | 9.19E-01 |
| | | | | | | | | | | | | Interaction model | 1.14336845 | 0.3433103 | 3.8079 | 8.27E-01 |
| Psychiatric Disorders | Tamoxifen | CYP2D6 | CYP2D6*10 (rs1065852) | T | Dominant | Depression | Pre- , Peri- & Post-menopausal | Al-Mamun 2017 | Age, BMI | OR=2.15E+08, p=0.0140 | Age, BMI, assessment centre, first five principal | Main effects model | 1.02205069 | 0.9755618 | 1.07075 | 3.58E-01 |
| | | | | | | | | | | | | Interaction model | 1.397593 | 0.7752137 | 2.51965 | 2.70E-01 |
| Psychiatric Disorders | Tamoxifen | CYP2D6 | CYP2D6*10 (rs1065852) | T | Dominant | Depression | Pre- , Peri- & Post-menopausal | Al-Mamun 2017 | Unadjusted | OR=33.5350 (2.0359, 552.3914) | Assessment centre, first five principal components | Main effects model | 1.01838464 | 0.9723326 | 1.06662 | 4.40E-01 |
| | | | | | | | | | | | | Interaction model | 1.41390376 | 0.7850749 | 2.54641 | 2.49E-01 |

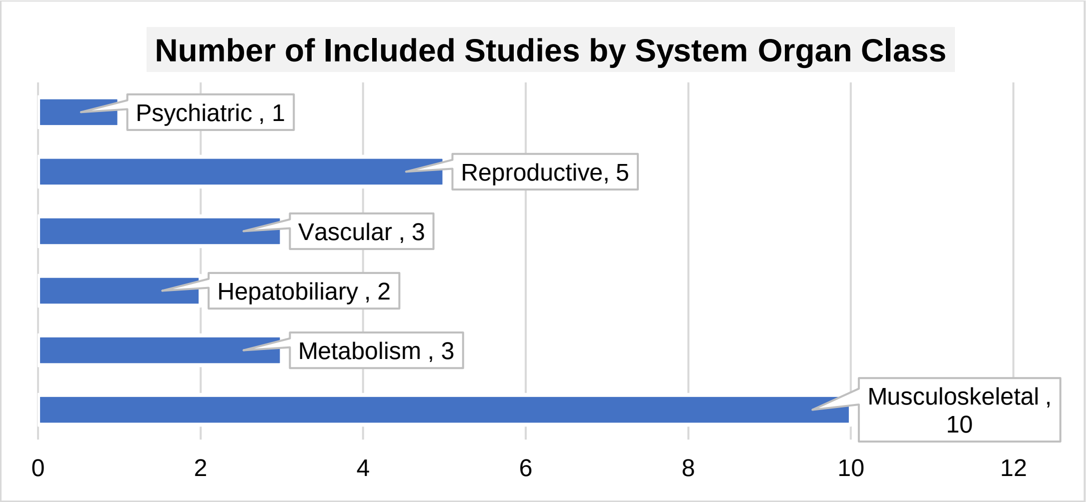

*Figure S1 Number of identified PGx studies of ADEs related to endocrine therapy by system organ class.*

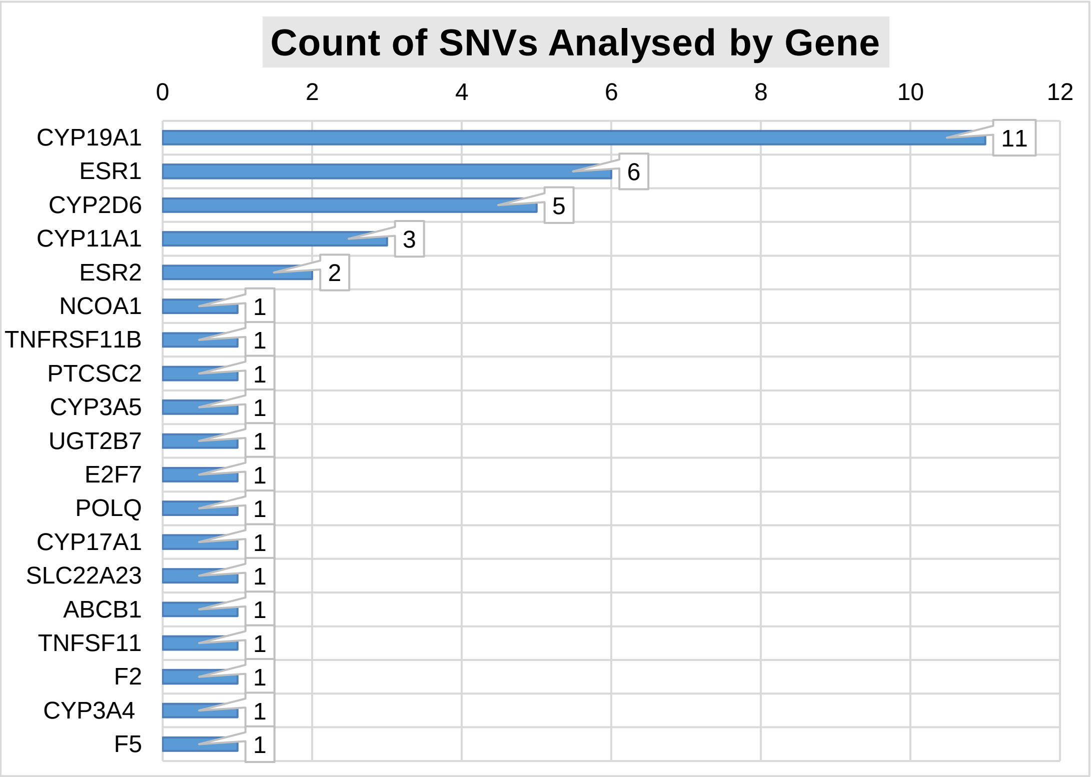

*Figure S2 Count of SNVs analysed in the UK Biobank per gene regarding PGx studies of ADEs related to endocrine therapy.*
